# Comprehensive cerebrospinal fluid analysis indicates key roles for B cells in multiple sclerosis

**DOI:** 10.1101/2025.01.02.24319302

**Authors:** Camila Fernández-Zapata, Carolin Otto, Gerardina Gallaccio, Qianlan Chen, Meng Wang, Burulça Uluvar, Matteo Teves, Claudia Samol, Maria Buthut, Fabian R. Bösl, Adeline Dehlinger, GueHo Jang, Christian Böttcher, Helena Radbruch, Josef Priller, Patrick Schindler, Catarina Raposo, Sven Shippling, Rosetta Pedotti, Desiree Kunkel, Maik Pietzner, Christiana Franke, Peter J. Oefner, Wolfram Gronwald, Harald Prüß, Johannes Lohmeier, Friedemann Paul, Klemens Ruprecht, Chotima Böttcher

**Affiliations:** Experimental and Clinical Research Center, a cooperation between the Max Delbrück Center for Molecular Medicine in the Helmholtz Association and Charité Universitätsmedizin Berlin, Germany; Max Delbrück Center for Molecular Medicine in the Helmholtz Association (MDC), Berlin, Germany; Department of Neurology and Experimental Neurology, Charité-Universitätsmedizin Berlin, Berlin, Germany; Neuroscience Research Center, Charité-Universitätsmedizin Berlin, Berlin, Germany; Computational Medicine, Berlin Institute of Health at Charité-Universitätsmedizin Berlin, Berlin, Germany; Institute of Functional Genomics, University of Regensburg, Am Biopark 9, 93053 Regensburg, Germany; Department of Psychiatry and Neurosciences, Clinical Neurotechnology Lab, Neuroscience Research Center, Charité - Universitätsmedizin Berlin, Berlin, Germany; Department of Neuropathology, Charité-Universitätsmedizin Berlin, corporate member of Freie Universität Berlin and Humboldt-Universität zu Berlin, 10117, Berlin, Germany; Neuropsychiatry and Laboratory of Molecular Psychiatry, Charité – Universitätsmedizin Berlin, corporate member of Freie Universität Berlin and Humboldt-Universität zu Berlin, Berlin, Germany; German Center for Neurodegenerative Diseases (DZNE), Berlin, Germany; Department of Psychiatry and Psychotherapy, School of Medicine and Health, Technical University of Munich and German Center for Mental Health (DZPG), Munich, Germany; University of Edinburgh and UK DRI, Edinburgh, UK; F. Hoffmann-La Roche Ltd., 4070 Basel, Switzerland; Flow & MassCytometry Core Facility, Berlin Institute of Health at Charité - Universitätsmedizin Berlin, Berlin 13353, Germany; Precision Healthcare University Research Institute, Queen Mary University of London, London, UK; MRC Epidemiology Unit, University of Cambridge School of Clinical Medicine, Institute of Metabolic Science, Cambridge, UK; Institute of Neuroradiology, Charité-Universitätsmedizin Berlin, Berlin, Germany

## Abstract

Multiple sclerosis (MS) is a complex inflammatory and neurodegenerative disease of the central nervous system (CNS) with a multifaceted pathophysiology, likely involving a variety of mechanisms and effectors. To characterize the spectrum of cellular and molecular factors involved in MS at an unprecedented level, we here performed a comprehensive analysis of cerebrospinal fluid (CSF) and peripheral blood using multiple high-dimensional technologies, including mass cytometry, metabolomics and proteomics (NULISA and Olink Explore^®^ 3072). Enriched B cells and proteins involved in B cell functions in the CSF separated MS patients from other neurological disease entities. Specific B cell subpopulations and molecular markers including gut-microbiota-derived metabolites and neurofilament light protein, a marker of neuroaxonal damage, in CSF correlated with clinical (acute vs. stable disease) and/or radiological (gadolinium enhancement) disease activity. Altogether, unbiased broad phenotyping suggests key roles of diverse B subpopulations and B cell related molecular markers in MS, which are associated with both, inflammatory and degenerative aspects of the disease and may serve as disease activity and treatment response biomarkers.

---

Multiple Sclerosis (MS) is a chronic, immune-mediated, inflammatory and neurodegenerative disease of the central nervous system (CNS). It is the most common neurological disease leading to permanent disability in young adults. MS exhibits an extraordinary heterogeneity of clinical features, genetic and environmental risk factors, and associated mechanisms of tissue damage.^1^ While the exact pathomechanisms of MS remain elusive, they comprise both inflammatory and degenerative processes.^2^ One of the fundamental pathomechanisms involves cell-mediated and humoral immune dysregulation, which is likely triggered and sustained by both genetic as well as non-genetic and environmental risk factors.^1,3–8^ In particular, B cells have emerged as crucial contributors to the immunopathology of MS. High efficacy of CD20 depleting therapies, such as ocrelizumab, has put the involvement of B cells in MS pathology in the spotlight.^9^ The presence of B cells in the cerebrospinal fluid (CSF) of MS patients is well-documented,^10–14^ and they are thought to drive ongoing CNS damage, contributing to the progression of disability.^15^ Despite the success of therapies like ocrelizumab in reducing relapses, benefits against underlying disease progression are only partial.^15^ This suggests that a more detailed characterization of the B cell subsets in the CSF is needed. Mapping the repertoire of B cell subtypes in MS patients may not only reveal additional therapeutic targets, but also provide valuable insights into disease activity and progression. These insights could potentially be linked to different B cell subsets in distinct ways. By identifying specific ‘good or bad’ B cell subpopulations, it may become possible to refine therapeutic strategies and develop biomarkers that aid in diagnosing and monitoring disease progression.

One of the recommended pathways to cures for MS research roadmap (i.e., the stop pathway)^16^ suggests dedicating efforts to early personalized diagnosis and therapy to minimize CNS damage and delay the accumulation of disability.^17,18^ To do so, precision medicine diagnostics using multimodal assessments is the most promising tool. In this study, we combine multiple state-of-the-art technologies to holistically identify cellular and molecular features in the CSF and peripheral blood that are associated with MS and its disease activity. Specifically, we identify enriched B cells and proteins involved in B cell activation in the CSF of MS patients, while subtle differences in T cells and myeloid cell populations are found in CSF and the peripheral blood. We further identify memory and antibody secreting B cells (ASCs), as well as proteo-metabolomic markers including aryl hydrocarbon receptor (AhR) ligands that associate with clinical disease activity and/or radiographic disease activity in patients with MS (pwMS). Our findings emphasize the key role of B cells and related proteo-metabolomic factors in MS and suggest distinct roles of different B cell subsets.

## Results

### Cellular signature in CSF distinguishes MS patients from other neurological diseases

To comprehensive characterize effector factors involved in MS, we performed holistic analyses of CSF and peripheral blood in a total of 66 pwMS and 128 non-MS patients ranging from Alzheimer’s disease (n=46), dementia and cognitive impairment (n=24), long-COVID-19 (n=24), neurosyphilis (n=2) to other neurological diseases (n=32) (Fig. 1a, Supplementary Table 1 and 2, see also Patients’ characterization in ONLINE METHODS).

**Fig. 1.**
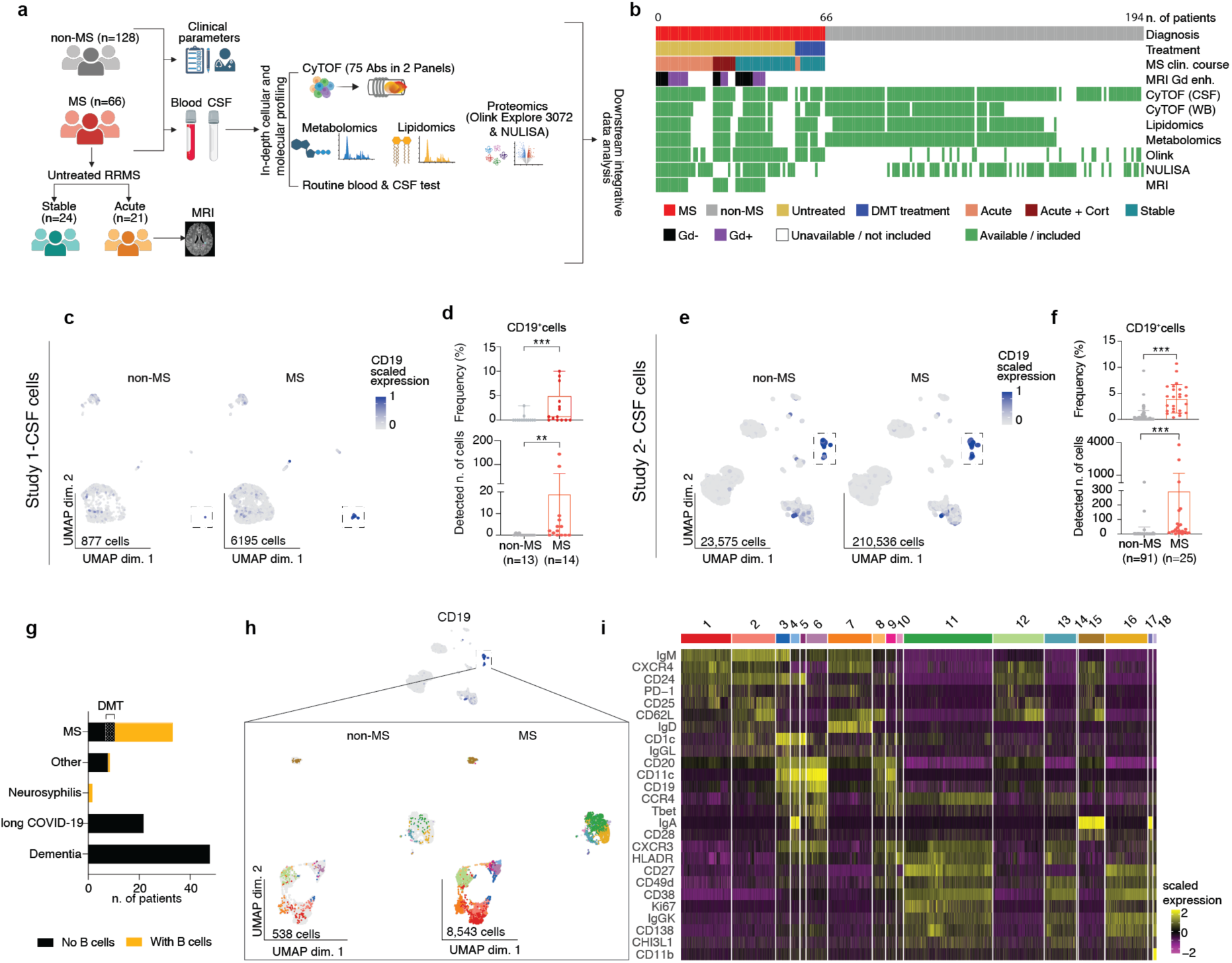
Multimodal assessment of whole blood and CSF samples from MS and non-MS patients. **a,** Schematic depiction of the experimental workflow including recruitment of MS and non-MS donors, inclusion of clinical parameters, and collection of blood and CSF samples. Immune cells from whole blood and CSF were used for immune profiling using CyTOF with two antibody panels. The remaining CSF and plasma were used for targeted metabolomics, lipidomics and proteomics using the Olink Explore and NULISA platforms, as well as for routine blood and CSF tests. All data was then analyzed and integrated. **b,** Map of samples included in the study, stratified by diagnosis, disease modifying treatment (DMT) at time of sample collection, MS clinical course and presence of Gd enhanced lesions in the MRI scans. Data availability from each experimental platform is also included. **c, e,** Two-dimensional uniform manifold approximation and projection (UMAP) plots colored by CD19 expression in cells from the CSF of non-MS and MS patients included in Study 1 and Study 2 respectively. Each dot represents one cell. **d, f,** Bar plots showing frequency (%) and detected number of CD19^+^ cells in the CSF of non-MS and MS patients included in Study 1 and Study 2 respectively. **g,** Bar plot showing distribution of patients with detected presence of B cells in the CSF. **h,** UMAP plots showing unsupervised clustering of B cells present in the CSF of non-MS and MS donors in Study 2, colored by subcluster ID. **i,** Phenotypic heatmap of B cell subclusters, with rows representing different markers and columns corresponding to individual cells.

Utilizing multi-omics and high-dimensional technologies, we obtained comprehensive profiles of cells, proteins, metabolites and lipids in CSF and paired whole blood (WB)/plasma samples (Fig. 1a,b). First, we conducted a preliminary mass cytometry (CyTOF) analysis of CSF samples from a small cohort of patients (Study 1: 14 untreated MS patients and 13 non-MS patients). Using antibody Panel A (37 antibodies, Supplementary Table 3), specifically designed for B cell phenotype characterization, we detected a significant enrichment of CD19^+^ B cells in CSF of pwMS (Fig. 1c,d). This finding was subsequently validated in a larger patient cohort (Study 2), encompassing 34 pwMS and 81 patients with non-MS diseases (Fig. 1e,f). For study 2, we additionally employed Antibody Panel B (37 antibodies, Supplementary Table 4) to analyze a wider spectrum of immune cell populations. Panel B enabled the characterization of CD3^+^ T cells and CD3^-^ cells, with the latter populations comprising myeloid cells, natural killer (NK) cells, and B cells (collectively termed MNB cells). This comprehensive approach, combining Panel A and B, facilitated a thorough examination of diverse immune cell subsets within the samples simultaneously (Supplementary Table 5 and 6, and Extended Data Fig. 1 for CSF, and Extended Data Fig. 2 and 3 for whole blood). Interestingly, among patients with non-MS diseases, we detected B cells in CSF of only two patients, both with neurosyphilis (Fig. 1e-g), in line with previous reports^19,20^, but in none of the remaining patients (Fig. 1g). Of note, these results were derived from the analysis of 3 ml CSF samples. This relatively small volume underscores the sensitivity of our analytical methods and highlights the differential B cell phenotypes present in the CSF of pwMS, as compared to other non-MS patients. Based on marker expressions, B cells in the CSF could be characterized into 18 different clusters (B1-B18) including CD25^+^CD62L^+^ naïve/regulatory B cells (B1, B2, B7, B12 and B15), CD11c^+^ atypical memory B cells (B3, B4, B5, B6, B8 and B9), and antibody secreting cells (ASCs) (B10, B11, B13, B16, B17 and B18) (Fig. 1h,i).

We also confirmed the increase in the presence of CSF B cells in pwMS (in Study 2) with the second antibody panel (Panel B: CD19^+^ clusters MNB1, MNB6 and MNB9, Extended Data Fig. 4a-c). In addition, a CD141^+^CD68^+^ myeloid cell cluster (MNB8) and two T cell clusters (CD4^+^ICOS^+^CD226^+^ T follicular helper (Tfh) cells (T5) and CD4^-^CD8^-^TIGIT^+^ double negative (DN) T cells (T8) with possible regulatory phenotype) were also detected at higher frequency in the CSF of MS patients (Extended Data Fig. 4a-c). In whole blood, the most prominent changes included a decreased proportion of diverse myeloid cell populations, which corresponded with an increased frequency of classical monocytes (M3) in MS patients (Extended Data Fig. 4a,d,e).

### CSF protein profiles underscore the key roles of B cells in MS

To identify soluble proteins associated with B cell enhancement in the CSF of pwMS, we next selected a subset of treatment-naïve MS patients (from both study 1 and study 2) and performed high-dimensional protein analysis using nucleic acid linked immuno-sandwich assay (NULISA) platform, which can detect up to 370 protein markers, in the CSF samples from patients with and without MS.^21^ Comparative analysis revealed 24 differentially expressed proteins (DEPs), with 21 of these showing increased levels in untreated pwMS (Supplementary Table 7). Among them were CD27, transmembrane activator and cyclophilin ligand interactor (TACI or TNFRSF13B), IFNγ, lymphotoxin alpha (LTA), IL12β, IL-10, TNF and CXCL13. These proteins are predominantly associated with B cell maturation, activation and immune response regulation (Fig. 2a,b). This finding suggests a distinct proteomic profile in the CSF of MS patients, which is linked to B cell enrichment in the CSF as shown in Fig. 1. Interestingly, among the DEPs increased in MS were also two proteins involved in neuronal signaling, brain acid soluble protein 1 (BASP1) and TAR DNA-binding protein 43 (pTDP43-409). Accumulation of TDP-43 has been associated with several neurodegenerative diseases, though to our knowledge, not with MS. Furthermore, three proteins were found to be decreased in MS as compared to non-MS patients, these were p-Tau181, p-Tau231 and IL7, increase of p-Tau in the non-MS group is most likely due to the inclusion of AD patients in this group. Protein interaction analysis from the STRING database visualized using Cytoscape^22^ revealed the functional network between differentially expressed proteins especially with IFNγ, IL-10, CSF2 (or GM-CSF), CD27, TNF, CXCL13, CD4 and CTLA4 (Fig. 2c).

**Fig. 2.**
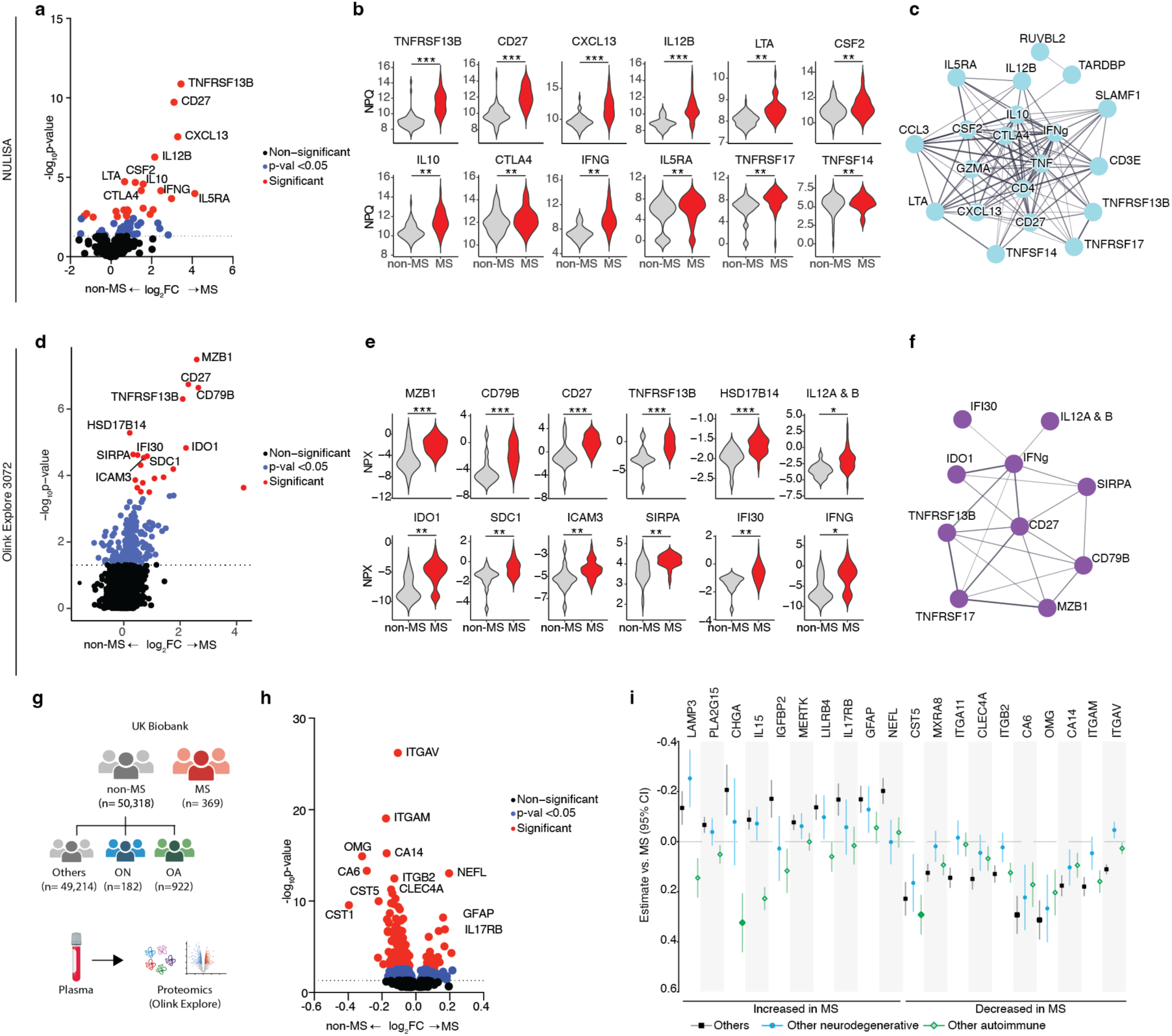
CSF protein profiles distinguish MS patients from non-MS patients. **a, d,** Volcano plot showing the differentially expressed proteins (DEPs) in CSF between non-MS and MS patients from NULISA and Olink Explore platforms respectively. Calculated using linear mixed effects model adjusted for age. Proteins with statistically significant difference between groups after correction for multiple testing (Benjamini & Hochberg) are colored in red; proteins with *p*-value <0.05 colored in blue, others are in black. Each dot represents one protein, and horizontal dashed line represents a *p-* value threshold of 0.05. **b, e,** NULISA Protein Quantification (NPQ) and normalized protein expression (NPX) of the top 10 DEPs between non-MS and MS patients for NULISA and Olink experiments respectively. **c, f,** Protein interaction networks from the DEPs obtained with STRING database in Cytoscape, only proteins with interactions are shown. **g,** Olink plasma protein profiles from the UK Biobank were used to test differences between MS patients and non-MS, including patients with other neurological (ON) and other autoimmune (OA) diseases. **h,** Volcano plot showing DEPs between MS and non-MS patients, calculated using linear regression adjusted for age, sex, and other technical covariates. Proteins with statistically significant difference between groups after Bonferroni correction for multiple testing are shown in red; proteins with *p*-value <0.05 colored in blue, and non-significant proteins are in black. Each dot represents one protein, and the horizontal dashed line represents a *p-* value threshold of 0.05. **i,** Forest plot with beta coefficients and their 95% confidence intervals of plasma protein levels (NPX values), showing proteins with most elevated and reduced levels in the MS group as compared to those with other neurodegenerative disorders, other autoimmune disorders and controls. Testing was conducted using a linear regression model for each protein assay, adjusting for sex, age and additional technical covariates. \**p* < 0.05, \*\**p* < 0.01, \*\*\**p* < 0.001.

To further explore in-depth protein profiles, we performed a large-scale CSF proteomics using the Olink Explore 3072 platform^23^, which can detect 3072 different proteins. We detected 20 DEPs, all significantly increased in the CSF of MS patients as compared with non-MS patients (Fig. 2d, Supplementary Table 8). Four of these proteins had also been detected by NULISA (i.e., CD27, TNFRSF13B, IL12B and IFNγ). The detected DEPs are involved in immune cell responses and B cell development and function such as marginal zone B cell-specific protein (MZB1), CD79B, CD27, TNFRSF13B, syndecan 1 (SDC1 or CD138), intercellular adhesion molecule 3 (ICAM3), signal regulatory protein alpha (SIRPα), IL12β and gamma-interferon-inducible lysosomal thiol reductase (GILT or IFI30), as well as IFNγ, enzymes involved in metabolic pathways such as hydroxysteroid 17-beta dehydrogenase 14 (HSD17B14) and indoleamine 2,3-dioxygenase 1 (IDO1) (Fig. 2e). Among these proteins, we found functional networks between IFNγ and IFI30, IL12A, CD27, IDO1, SIRPα, TNFRSF13B and TNFRSF17 as well as between CD27 and CD79b, MZB1, TNFRSF17 (or B-cell maturation antigen, BCMA), TNFRSF13B, IDO1, SIRPα and IFNγ as by protein interaction analysis using Cytoscape (Fig. 2f). Using this comprehensive set of DEPs (from either NULISA or Olink), patients with MS could be accurately discriminated from non-MS patients (Extended Data Fig. 4f,i). These identified biomarker proteins (both from Olink and NULISA) aligned with the CSF biomarkers defined in a previous study by Åkesson et al.^24^, which was conducted on a limited cohort of pwMS and healthy controls. In that study, the CSF and plasma levels of neurofilament light chain (NfL, or NEFL), a well-studied biomarker for neuro-axonal injury, were proposed as one of potential predictors of age-related MS score and, consequently, disability worsening. However, in our current study, the CSF levels of NfL (determined by both NULISA and Olink) showed no significant difference between pwMS and non-MS patients, after age-correction (Extended Data Fig. 4g,h,j,k).

When testing the diagnostic potential of NfL in plasma from the UK biobank cohort (Fig. 2g; non-MS: n = 50,318; MS: n = 369), we detected differentially expressed plasma proteins in MS participants including elevated plasma NfL (Fig. 2h,i, Supplementary Table 9). Nevertheless, plasma NfL levels were comparable between MS participants and those with other autoimmune or neurological diseases (Fig. 2i). This suggests that NfL is more indicative of neuro-axonal injury and/or disease activity rather than a specific diagnostic biomarker for distinguishing MS from other neurological diseases.

Despite less abundance of potential plasma biomarkers in pwMS, our previous study conducted on a cohort of pwMS treated with ocrelizumab (n = 29, Extended Data Fig. 5a), an anti-CD20 antibody confirmed a reduction in B cell proportion in peripheral blood, accompanied by decreased plasma NfL levels (Extended Data Fig. 5b,c). Additionally, we detected diminished levels of plasma biomarkers associated with B cell activation and involved in MS pathology (as shown in Fig. 2) such as TNFRSF13B, TNFRSF13C, CXCL13, TNFRSF9 and TNFSF9 (Extended Data Fig. 5d, Supplementary Table 10). Of note, these B cell biomarkers were detected increased in CSF of pwMS as shown in Fig. 2, as compared with non-MS patients. These markers showed high interaction (Extended Data Fig. 5e) and significant correlation with B cell proportion in the peripheral blood (Extended Data Fig. 5f). Since ocrelizumab is widely recognized as a potent drug that effectively reduces relapse episodes in RRMS, these B cell markers, in conjunction with NfL, may serve as valuable predictors for monitoring treatment efficacy.

### Antibody secreting cells in the CSF of pwMS correlate with biomarker proteins and clinical disease activity

To better understand the B cell composition in the CSF of untreated pwMS (Supplementary Table 2), we conducted a second round of cell-phenotype clustering using PCA. This unsupervised clustering of CyTOF data (Fig. 1h and i) revealed three main subsets of B cells, identified as CD20^+^CD27^+^CXCR3^hi^CD11c^+^Tbet^+^ atypical/memory B cells, CXCR3^lo^CD20^+^CXCR4^+^CD62L^+^ CD25^lo^ naïve/regulatory B cells and CXCR3^+^CD20^lo/-^ CD38^+^CD27^+^CD138^lo/+^CHI3L1^lo^ antibody secreting cells (ASCs). The latter included pre-plasmablasts, IgA^+^/IgG^+^ plasmablasts and plasma cells (Fig. 3a-d). Interestingly, the CSF levels of the DEPs detected by Olink and NULISA (as shown in Fig. 2) showed correlations mainly with the frequency of IgA^+^ plasmablast (B17) and other ASCs in CSF, whereas only IFNG and IL12A&B showed an association with memory and naïve/regulatory B cells (Fig. 3e, Supplementary Table 11 (NULISA) and Supplementary Table 12 (Olink)).

**Fig. 3.**
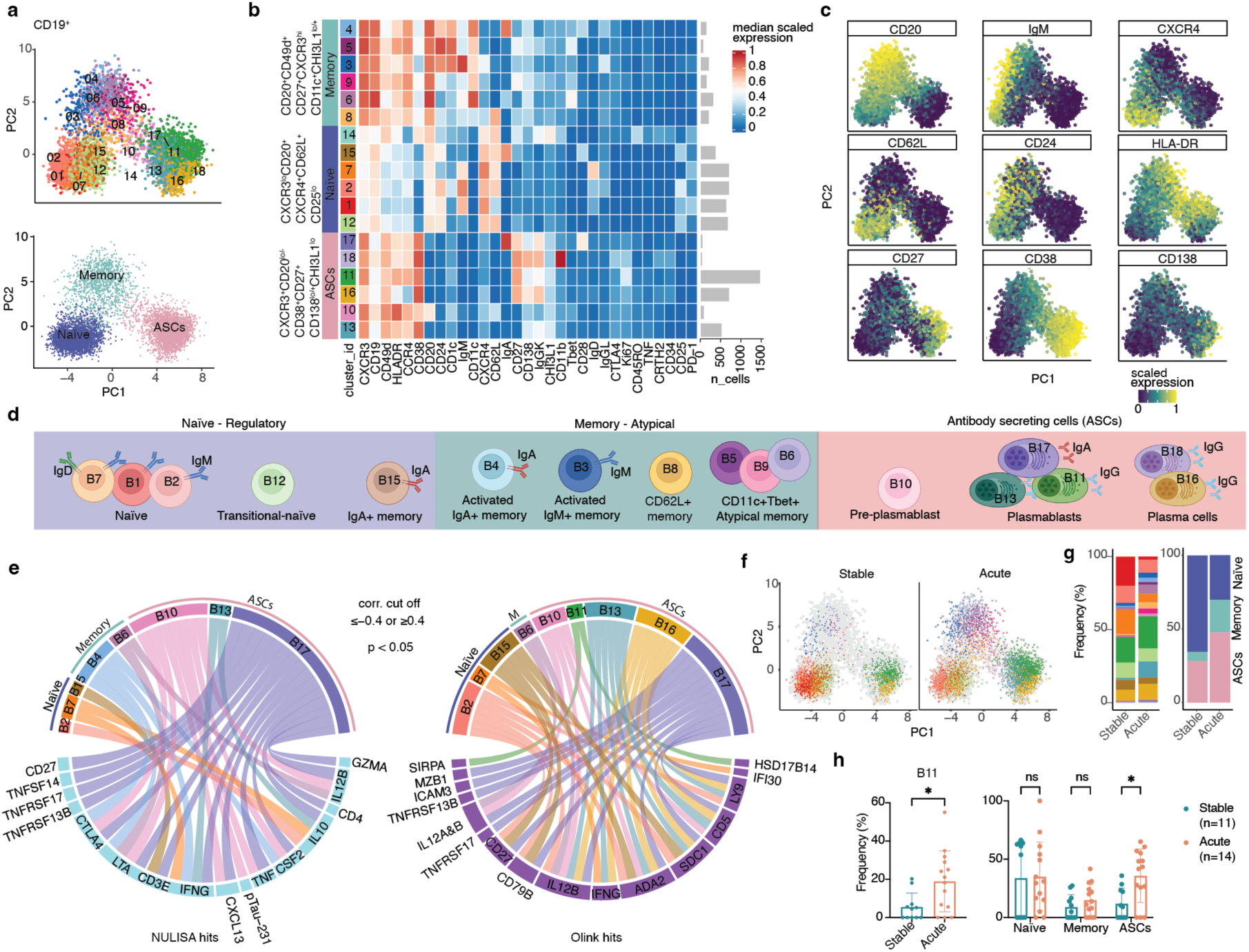
In depth characterization of B cells in the CSF of untreated MS patients. **a,** PCA plots showing CD19^+^ cells present in the CSF of untreated MS patients. The upper PCA plot is colored by subcluster ID, and the lower PCA plot is colored by major B cell categories. Each dot represents one cell. **b,** Heatmap showing median scaled expression from all markers for each subcluster, organized by the three major B cell categories. **c,** PCA plots colored by scaled expression of selected markers. **d,** Schematic illustration of B cell subclusters present in the CSF of untreated MS patients, each cell is colored according to the subcluster ID. Background colors correspond to the major B cell categories. **e,** Correlations between CSF B cell frequencies and DEPs from Olink and NULISA experiments are visualized in a chord diagram. Only correlations with a correlation coefficient ≤ -0.4 or ≤0.4 and *p*-value <0.05 are shown. **f,** PCA plots colored by subcluster ID showing cells present in MS patients with stable and acute clinical course. **g,** Frequency distribution of B cell subclusters and the three main B cell categories between stable and acute MS patients. **h,** Bar plots displaying frequency of B cell subcluster B11 and of the three main B cell categories. Statistical analysis using two-tailed Mann–Whitney U-test. \**p* < 0.05, \*\**p* < 0.01, \*\*\**p* < 0.001.

To determine if these B cell subsets and B cell biomarker proteins showed an association with MS clinical disease activity, we categorized untreated MS patients into acute (experiencing clinical relapse within 30 days before CSF and blood collection) and stable (no relapse for at least 30 days before sample collection). Olink and NULISA analyses did not detect CSF and plasma proteins that were significantly different between patients with and without acute relapse (Extended Data Fig. 6a-c). However, when comparing the B cell composition of acute patients with those of stable patients, the proportion of memory B cells and ASCs tended to be higher in acute patients (Fig. 3f-h), but only Ki67^+^IgGK^+^ plasmablast (B11) differed significantly between these two groups (Fig. 3h).

In addition, using the antibody panel B, the frequency of both, CD95^+^CD11c^+^CD69^+^ activated B cells (MNB1) and CD4^+^TIGIT^+^ regulatory T cells (T5), was increased in CSF of MS patients with acute relapse, whereas the frequency of CD4^+^CD161^+^ memory T cells (T1) in CSF was lower during relapse phase (Extended Data Fig. 6d,e). In contrast to CSF, only two small Tbet^+^ B cell subsets differed in peripheral blood of MS patients with acute versus stable disease course (Extended Data Fig. 6f,g).

### Immuno-proteo-metabolomic markers in CSF of pwMS correlate with clinical disease activity

Next, we determined whether, in addition to immune-proteomic profiles, metabolomic features in CSF and plasma of pwMS will be distinct from those of non-MS patients. We performed metabolomic and lipidomic profiling including tryptophan-derived metabolites using mass spectrometry and nuclear magnetic resonance (NMR) (see details in ONLINE METHOD section). When comparing pwMS with non-MS patients, only minor increases of CSF levels of pyroglutamate and threonine could be detected in non-MS patients (Extended Data Fig. 7a), whereas no differences in tryptophan metabolites and lipidomic profiles could be observed between non-MS patients and pwMS(Extended Data Fig. 7b,c).

Among untreated pwMS, we also detected a significant (p<0.01) lower levels of pyroglutamate in the CSF of acute relapse patients, while the plasma level of phenylalanine was increased (Fig. 4a,b). Levels of tryptophan metabolites were also found to differ between patients with acute relapse and stable disease (Fig. 4a,b). The CSF level of 5-hydroxyindole-3-acetic acid (HIAA), the primary metabolite of serotonin, was significantly (p<0.05) higher in the former, whereas plasma levels of indole acetic acid (IAA) and indole lactic acid (ILA) were higher in the latter.

**Fig. 4.**
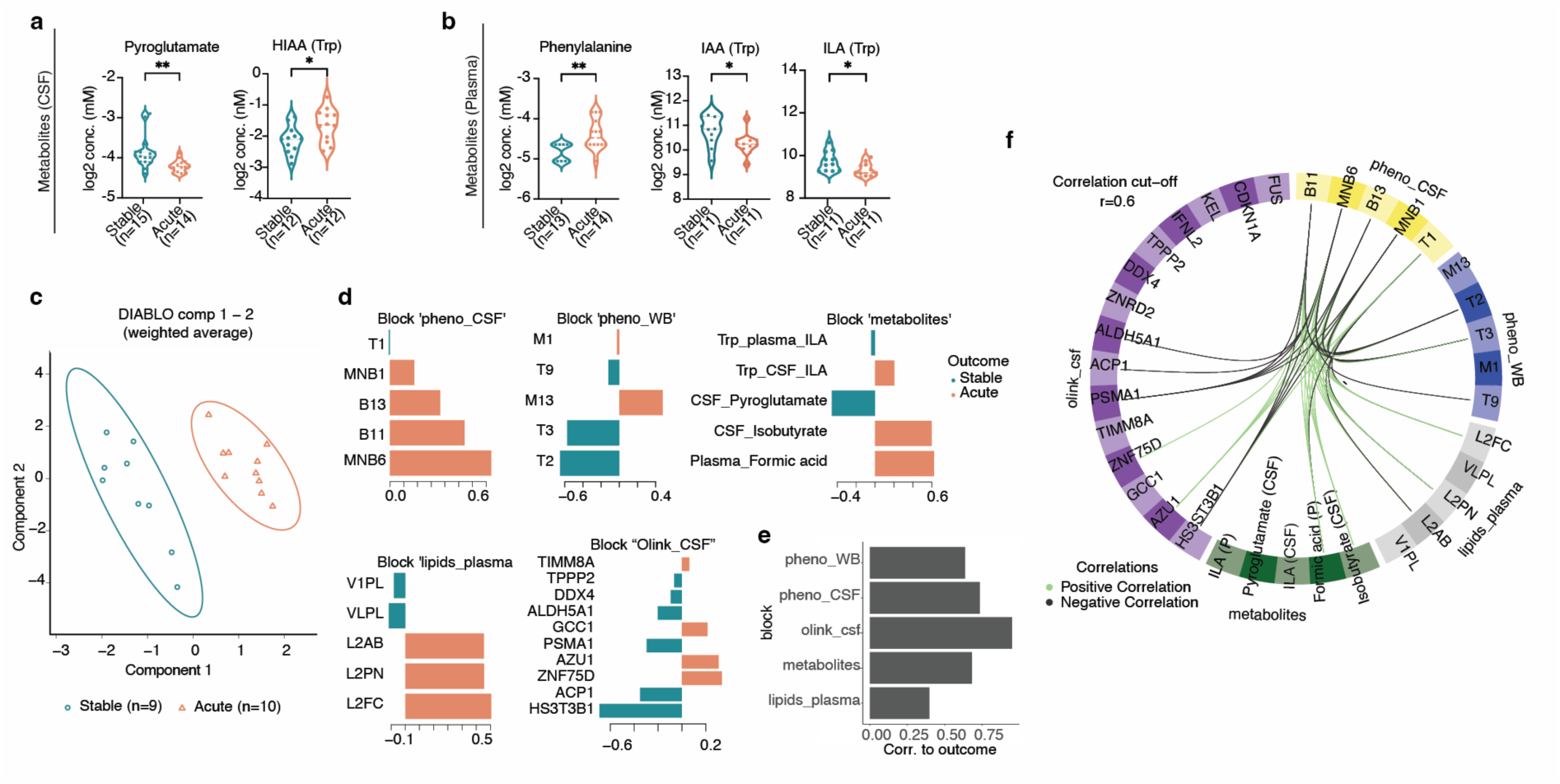
Multimodal biomarkers distinguish MS patients with acute clinical relapse. **a, b,** Violin plots of relevant metabolites with different concentration levels in CSF and plasma respectively. Statistical analysis using two-tailed Mann–Whitney U-test. **c,** Samples are plotted in the weighted two-component space obtained after performing supervised discriminant analysis with multiblock partial least square analysis (blockPLSDA or DIABLO). **d,** Loading plot for the variables selected by DIABLO on component 1. Variables are organized bottom to top according to decreasing coefficients and colored by group in which median expression value is higher. **e,** Bar plot showing correlation to outcome from each of the blocks or datasets used in DIABLO. **f,** Circus plot showing interactions above a 0.6 threshold between all blocks used for DIABLO analysis. Internal links in green show positive correlations while black links show negative correlations. Variables on side quadrants are colored according to their block of origin. \**p* < 0.05, \*\**p* < 0.01, \*\*\**p* < 0.001.

Next, we used DIABLO (Data Integration Analysis for Biomarker Discovery using Latent variable approaches for Omics studies, implemented in the R package mixOmics^25^) on paired high-dimensional datasets (CyTOF, Olink, lipidomics and metabolomics) to construct a supervised machine-learning classifier for the distinction of acute and stable clinical courses in untreated pwMS (Fig. 4c-f). Of note, DIABLO focuses on maximizing the correlation between different datasets to identify key variables that discriminate between studied groups. It does not consider p-values during the feature selection, instead it utilizes a multivariate approach based on projection to latent structures to select variables according to their contribution to the model’s predictive performance and stability across cross-validation^25^. DIABLO identified comprehensive cellular and molecular markers that showed discriminative ability and correlation between technologies, and thus could differentiate between acute and stable clinical course in untreated pwMS (Fig. 4c). These included ASCs (B11 and B13) and CD95^+^ B cells (MNB1 and MNB6) in the CSF, and CD4^+^CD226^+^ICOS^+^ T cells (T2 and T3) as well as OPN^+^CD141^lo^ myeloid cells (M13) in the peripheral blood as well as soluble proteins (determined by Olink), metabolomic and lipidomic factors associated with inflammation and immune responses, such as isobutyrate, pyroglutamate and ILA as well as low density lipoprotein (LDL)-2 species (i.e., LDL-2 apolipoprotein B (L2AB), LDL-2 particle number (L2PN) and LDL-2 free cholesterol (L2FC)) (Fig. 4d). Interestingly, Olink protein markers that were negatively correlated with acute relapse are known to be involved in the regulation of immune and inflammatory responses, particularly in the context of anti-viral activity such as DDX4, ACP1 and HS3T3B1. In contrast, protein markers involved in an activation of immune cells such as AZU1 and GCC1 were more pronounced in the CSF of patients with acute relapse (see more details in Supplementary Table 13). Overall, the latent component from the Olink protein profiles in the CSF showed the highest weight for discriminating between patients with and without acute relapse, whereas lipid profiles had the least impact on discrimination of the clinical activity (Fig. 4e). When testing the correlation between CSF immune cells with other variables, we detected a negative correlation of blood T cells as well as HS3T3B1 (Heparan sulfate glucosamine 3-O-sulfotransferase 3B1) and PSMA1 (proteasome subunit alpha type-1) with those B cell subsets detected at a higher proportion in CSF of pwMS with acute relapse, suggesting regulatory roles of these markers (Fig. 4f). In contrast, LDL-2 species (L2AB, L2PN and L2FC) showed positive correlation with CSF B cell subsets enhancing in acute relapse (Fig. 4f).

### Differential cellular and molecular profiles associated with Gadolinium enhancement

Gadolinium-based contrast agents are capable of highlighting areas of blood-brain barrier disruption (i.e., a sign of active neuroinflammation)^26^. Gadolinium-enhanced (Gd+) MR imaging is a well-known sensitive method for detecting active lesions in MS. However, based on observations in clinical practice, the correlation between contrast-enhancing lesions and clinical relapses or disability is modest. Gd-positive lesions indicate active neuroinflammation, but not all patients showing clinically active disease course show Gd enhancement.^27–29^ Differential pathways or biomarkers that distinguish between clinical and radiographic disease activity remain to be investigated to help physicians in making treatment decisions in pwMS. To explore differential cellular and molecular biomarkers in treatment-naïve patients with radiographic disease activity compared with those who have clinical relapses, we normalized available MRI data obtained from routine diagnosis (Gd-: n = 12; Gd+: n = 13) (Supplementary Table 14 and Fig. 5a), and categorized untreated MS patients into groups with Gd-enhancing lesions (Gd+) and those without (Gd-), further distinguishing between those with (acute) and without (stable) clinical relapse (Fig. 5b). We consistently detected a higher proportion of memory B cells and ASCs in patients with clinical relapse (either Gd+ or Gd-) as compared with stable patients (either Gd+ or Gd-) (Fig. 5c).

**Fig. 5.**
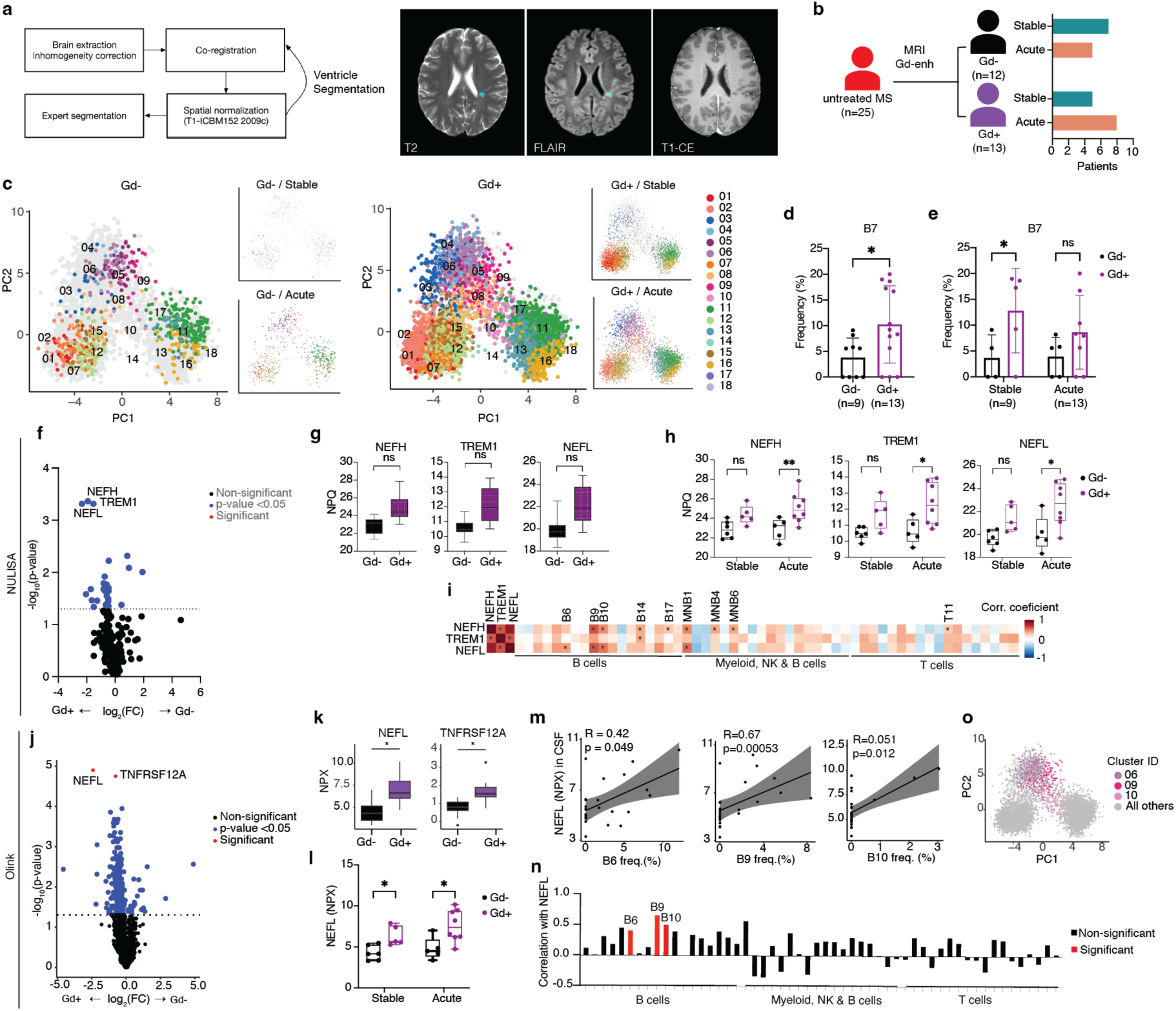
Immune and molecular signatures in MS patients with and without Gd enhanced lesions in MRI. **a,** Workflow used for analysis of MRI data. **b,** Distribution of untreated MS patients. **c,** PCA plots of B cells present in the CSF of patients with (Gd+) and without (Gd-) Gd-enhancement and corresponding stable and acute B cell populations. **d,** Distribution of B cell subclusters and the three major B cell populations between Gd- and Gd+ patients. **e,** Bar plots showing differences in frequency of CSF immune cell populations. Statistical analysis using two-tailed Mann–Whitney U-test. **f, j,** Volcano plots showing the DEPs in the CSF of patients with and without Gd-enhancement for NULISA and Olink experiments respectively, calculated using a two-way Mann Whitney U test. Proteins with statistically significant differences between groups after correction for multiple testing (Benjamini & Hochberg) are colored in red; those with a *p*-value <0.05 are colored in blue, others are in black. Each dot represents one protein and horizontal dashed line represents a *p*-value threshold of 0.05. **g, h, k, l,** NPQ and NPX levels for the DEPs. **j, m, n,** Correlations between NEFH, TREM1 and NEFL levels (NPQ) or NEFL levels (NPX) and frequency of immune cells present in the CSF. In (n) significant correlations are colored in red; others are in black. Detailed correlation plots of significant ones in (m). o, PCA of B cells colored by subclustercluster identity of those significantly correlated populations, all others in grey. \**p* < 0.05, \*\**p* < 0.01, \*\*\**p* < 0.001.

Unlike clinical relapses (as shown in Fig. 3f-h), we detected significant differences in the proportion of CD62L^+^CXCR4^+^ naïve B cells (B7, characterized by using antibody panel A, Fig. 5d) in the CSF of Gd+ pwMS. This B cell subset was proportionally comparable between acute and stable disease course (Fig. 5e). In addition, subtle changes of CD95^+^CD11c^+^ B cells (MNB1) and CD4^+^CD8^+^ double positive T cells (T6) were also found in CSF of Gd+ pwMS (Extended Data Fig. 8a). Notably, we detected an elevated proportion of CD20^-^ CD19^lo^CD38^+^IgA^+^ B cells in the peripheral blood of Gd+ pwMS, accompanied by proportional changes in diverse myeloid cell populations, including granulocytes (Extended Data Fig. 8c,d). Comparing NULISA protein profiling between Gd- and Gd+ patients revealed a difference in the CSF level of neurofilament light chain (NEFL or NfL) (Supplementary Table 15 and Fig. 5f,g). But the difference reached statistical significance only in Gd+ patients with acute relapse (Fig. 5h), a trend that was replicated for neurofilament heavy chain (NEFH or NfH) and triggering receptor expressed on myeloid cells 1 (TREM1) (Fig. 5h). Of note, CSF levels of NfL, NfH and TREM1 were comparable between patients with and without clinical relapse (Fig. 5h, stable vs acute). Interestingly, CSF levels of NfL and NfH proteins showed a positive correlation with the proportion of several CSF immune populations, among them CD11c^+^ atypical memory B cells (B6 and B9) and ASCs (B10) (Fig. 5i, see also phenotypic map on Fig. 3b). The NfL findings could be confirmed using Olink-Explore 3072 analysis (Supplementary Table 16 and Fig. 5j-l). In addition, TNFRSF12A, a receptor for the cytokine TWEAK that is involved in MS neuroinflammation^30^, was also detected at a higher level in the CSF of Gd+ patients (Fig. 5j and k). Of note TNFRSF12A was not included in the NULISA panel. In line with the results obtained using NULISA, the CSF levels of Olink NfL showed significant correlation with the proportion of CD11c^+^ atypical memory B cells and ASCs (B6, B9 and B10, respectively) (Fig. 5m, n and o). Similar to CSF protein profiles, levels of plasma NfL and NfH, as well as plasma Fms Related Receptor Tyrosine Kinase 4 (FLT4), were found higher in Gd+ pwMS (Extended Data Fig 8e,f). Correlation analysis between radiographic variables and immune phenotypes, along with metabolomic profiles, revealed a strong association between CD24^+^CD25^+^ regulatory B cells in the CSF and enhanced volume and lesion count (Extended Data Fig. 8g). However, only a small association was observed between radiographic parameters and metabolomic profiles (Extended Data Fig. 8h). Our findings suggest different kinetics and/or mechanism between clinical and radiographic relapse in pwMS.

### Cortisone treatment does not target B cells

High-dose corticosteroid therapy is commonly used to manage relapses in MS. While it can accelerate recovery from acute relapses and alleviate symptoms, it does not prevent future relapses or modify the disease’s long-term trajectory^31,32^. DIABLO analysis of high-dimensional cellular and molecular profiles of CSF and peripheral blood samples revealed unique differences between cortisone-treated and untreated patients with acute relapse (Fig. 6a,b).

**Fig. 6.**
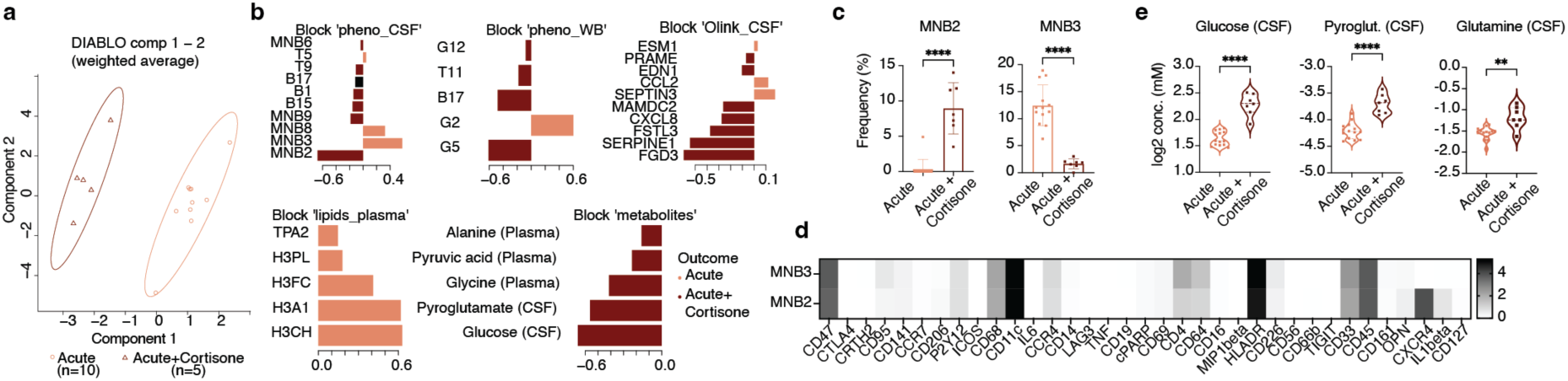
Multiomic signature of treated MS patients. a, Samples plotted in the weighted two-component space obtained after performing DIABLO to discriminate between acute and cortisone-treated acute (Acute+Cortisone) MS patients. **b,** Loading plots for the variables selected by DIABLO on component 1. Variables are organized bottom to top according to decreasing coefficients and colored by group in which median expression value is higher. **c,** Frequency of the top most contributing CSF immune populations and corresponding phenotypic heatmaps of normalized protein expression (**d**). **e,** Violin plots showing concentration levels of the two most contributing metabolites. \**p* < 0.05, \*\**p* < 0.01, \*\*\**p* < 0.001.

Among these changes in immune phenotypes, cortisone-treated patients showed a higher and lower proportion of CD33^+^CD11c^+^CXCR4^+^IL-1*β*^+^ myeloid cells (MNB2) and CD33^+^CD11c^+^CXCR4^-^ myeloid cells (MNB3) in the CSF, respectively, whereas B cell alterations were modest (Fig. 6b-d). In the peripheral blood, CD14^+^CD161^lo^ granulocytes (G2) were found at lower proportion in cortisone-treated pwMS, whereas CD14^-^CD161^+^ subset (G5) was at higher proportion in cortisone-treated pwMS (Fig. 6b). In line with previous studies, we observed a steady rise in CSF glucose and glutamine levels of cortisone-treated patients compared to untreated patients (Fig. 6b (metabolites) and e)^33^. In addition, we also detected an increased CSF levels of pyroglutamic acid under cortisone treatment (Fig. 6e), the metabolite which was also found to be reduced in patients with acute relapse as compared to stable patients (as shown in Fig. 4a). Most DEPs in the CSF of cortisone-treated patients were involved in cell migration and inflammation (Fig. 6b (Olink) and Supplementary Table 17).

These defined proteins did not overlap with those defined proteins specific to MS pathology (as shown in Fig. 2d) or those associated with clinical relapse (as shown in Fig. 4d).

## Discussion

The goal of this study was to identify cellular and molecular markers associated with MS and its disease activity. It has long been proposed that B cells are highly linked to MS pathology.^15^ The established efficacy of B cell-depleting anti-CD20 (aCD20) monoclonal antibody therapies in pwMS highlights the significant role of B cells in MS pathophysiology.^34,35^ However, there is limited understanding of their diverse involvements (e.g., pathogenic versus regulatory function) in MS. It also remains unclear how to target pathogenic B cells for therapeutic purposes, beyond an unspecific B cell depletion. Here, we identified an elevated frequency of CSF B cells as a common cellular marker discriminating MS patients from other neurodegenerative and neuroinflammatory diseases, which is in line with previous studies.^10,11,36^ Extending previous studies, we comprehensively characterized diverse B cell subsets in the CSF of pwMS and their correlations with disease activity and molecular markers including lipids, metabolites and protein markers.

In contrast to the CSF compartment, we did not detect highly significant changes in the blood B cell populations of pwMS, compared with non-MS patients, underscored the concept of CNS compartmentalized inflammation in MS.^37,38^ The increased number of CSF B cells correlated with elevated levels of CSF proteins involved in immune responses, B cell signaling, survival/differentiation and activation. Some of these proteins such as MZB1, CXCL13, LTA, CD79B, CD27, TNFRSF13B, SDC1 and ICAM3 have been already suggested as markers predicting the severity of disability worsening.^24^ CD27^36,39^ and CD79b^40^ are linked to B cell activation, with CD79b being a crucial part of the B cell receptor (BCR) complex, increased levels of these markers may indicate B cell activation and engagement in immune responses associated with the disease. Notably, anti-CD79b therapies, which induce a state of B cell anergy rather than cell depletion (like anti-CD20 therapies), show promise in autoimmune diseases.^41^ CXCL13,^42^ a chemokine associated with B cell recruitment and the elevated expression of lymphotoxin-*α* (LTA),^43^ a protein expressed by B cells that plays a critical role in humoral immune responses and the regulation of inflammation.^44^ Interestingly, soluble MZB1,^45^ a protein expressed in the marginal zone that plays a crucial role in B cell biology (especially in plasma cell differentiation and antibody secretion), was also found to be upregulated in the CSF of pwMS. This upregulation positively correlated with the presence of ASCs in the CSF of pwMS. Additionally, we detected elevated levels of pro-inflammatory cytokines, including IFN-γ and IL-12B. IFN-γ, often found at increased levels in MS lesions, is recognized for its role in promoting B cell differentiation into T-bet-expressing memory B cells.^46^ However, in our study, we did not observe a correlation between IFN-γ levels and CD11c^+^T-bet^+^ atypical memory B cells, whereas IL-12B showed a positive correlation with this B cell subset. Previous research has shown that CD11c^+^ B cells overexpress pro-inflammatory genes, including IL-12 subunits.^47^

All of the aforementioned protein markers demonstrated a positive correlation with an increased proportion of memory B cells and ASCs in the CSF, particularly IgA^+^CD138^+^CD27^+^ plasmablasts (B17). This correlation suggests that these soluble protein markers contribute to shaping the environmental factors that support B cell survival, differentiation, and function within the CSF of pwMS. However, the origin of IgA^+^ B cells and these protein markers remains unclear and technically challenging. Previous reports suggested gut-microbiota-specific IgA^+^ cells as a systemic mediator during active neuroinflammation in MS.^48^ In a mouse model of chronic demyelination (i.e., experimental autoimmune encephalomyelitis, EAE), the IgA^+^ B cell population was demonstrated to mobilize from the gut and provide a protective role in suppressing neuroinflammation.^49^ Complementing published studies, we have identified additional IgA^+^ B cell subsets: IgA^+^CXCR4^+^CD62L^+^ regulatory (B15) and IgA^+^CD11c^+^CD24^+^ (B4) memory B cells (as illustrated in Fig. 1i and 3b), demonstrating a correlation with protein markers elevated in the CSF. However, these B cell populations did not show an association with clinical disease activity in our study, thus differential roles of these three IgA^+^ populations in MS, as well as the links between them remain to be investigated. Instead, we identified differences in the proportion of Ki67^+^IgG^+^ (B11) and Ki67^-/lo^IgG^+^ (B13) plasmablasts, as well as CD11c^+^CD95^+^CD69^+^ (MNB1) and CD11c^-^CD95^+^ (MNB6) memory B cells, between pwMS experiencing acute relapse compared to patients with stable disease. CD95^+^ B cells are integral to the regulation of immune responses, particularly in the context of autoimmune diseases, by mediating apoptosis essential for maintaining B cell tolerance,^50^ suggesting a regulatory function of these cells during clinical relapse. DIABLO analysis revealed reduced CSF levels of protein tyrosine phosphatase 1B (PTP1B, encoded by ACP1), a protein involved in the removal of autoreactive B cells.^51^ The analysis showed a negative correlation between PTP1B and the proportion of CD95^+^ B cells in the CSF, suggesting possible B cell dysfunction during clinical relapse. Furthermore, a reduced CSF level of HS3ST3B1, a protein with an anti- viral activity,^52^ was detected in association with clinical relapse and increased IgG^+^/CD95^+^ CSF B cells. This suggests an involvement of B cells in the dysregulation of antiviral mechanisms in MS, similar to what has been reported on T cell.^53^ However, it remains technically challenging to precisely prove the function of each B cell subset in the CSF in vivo. Previous studies have demonstrated that B cells, particularly plasmablasts and plasma cells in the CSF, can have both protective and pathogenic effects. These effects are attributed to their ability to produce IL-10, generate antibodies against microbes, and function as antigen presenting cells.^15,54–56^ In our study, we have comprehensively characterized diverse B cell subsets in CSF. Each of these subsets likely serves a distinct function; however, some may possibly be transient precursors to others.

In addition to protein and immune profiles, we identified a number of interesting metabolic differences in CSF and peripheral blood between pwMS with stable and acute disease, including effects of corticosteroid therapy. Corticosteroids have also been reported to inhibit at sub-micromolar levels the enzyme glutathione reductase, which catalyzes the conversion of oxidized glutathione (GSSG) to reducing glutathione (GSH) utilizing NADPH generated by the pentose phosphate pathway.^57^ Glutathione is a pivotal molecule in oxidative stress and hydrogen peroxide detoxification. Inhibition of glutathione reductase has been shown to cause not only the expected increase in intracellular GSSG, but it also increased significantly protein glutathionylation, i.e., the reversible attachment of glutathione to cysteine residues of target proteins.^58^ A study on neutrophils has shown that switching to a cyclic pentose phosphate pathway is a key feature of oxidative burst in activated neutrophils.^59^ This also explains why glutathione reductase-deficient neutrophils produce paradoxically far less reactive oxygen species upon ex vivo and in vivo activation despite the impaired regeneration of glutathione.^60^ It is generally accepted, that generation of oxidizing radicals is a critical component of the inflammatory processes involved in MS, which ultimately lead to irreversible tissue damage.^61^ Inflammation-associated oxidative burst in activated microglia and macrophages, induced by upregulation of NADPH oxidase expression, appears to play a key role in active MS lesions in relation to free radical-mediated tissue injury. Therefore, one of the beneficial effects of corticosteroids in the treatment active MS lesions may be attributed to their ability to dampen oxidative burst by inhibiting the activity of glutathione reductase. This may explain the observed increase in CSF levels of pyroglutamate upon treatment with corticosteroids (Fig. 6e), while its levels in untreated patients with acute disease were even lower than those of patients with stable disease (Fig. 4a). This is also in line with the observation by Kim et al.,^62^ who found significantly increased CSF levels of pyroglutamate only in relapsed MS patients that received corticosteroids. Metabolomics analysis also revealed a significant increase in CSF glutamine levels following corticosteroid therapy (as shown in Fig. 6e). It is known that MS can be characterized by excessive levels of glutamate, which lead to excitotoxicity – a major cause of axonal damage and oligodendrocyte death.^63^ Activated macrophages and microglia in MS lesions express high levels of glutaminase, an enzyme that converts glutamine to glutamate, and are associated with axonal damage.^64^ Collectively, our analyses demonstrated an additional mechanism of action for corticosteroids: reduced conversion of glutamine to glutamate, possibly via inhibition of glutaminase activity. Here, in pwMS experiencing acute relapse, we observed higher CSF levels of 5-hydroxyindoleacetic acid (5-HIAA), the main metabolite of serotonin and a biomarker for serotonin metabolism,^65^ suggesting dysregulation of serotonin metabolism during acute relapse. In addition to glutathione and serotonin metabolism, we also observed increase in plasma levels of phenylalanine in MS patients with acute clinical relapse, possibly due to an impairment of phenylalanine 4-hydroxylase (PAH).^66^ Gut microbial dysbiosis, including a depletion of lactobacilli and a reduction of its metabolite ILA, may play an important role in the pathogenesis of MS.^67^ Probiotics rich in lactobacilli have been shown to improve clinical symptoms in MS patients.^68,69^ Our study further supports the involvement of microbiota-derived tryptophan metabolites in modulating MS disease activity via the aryl hydrocarbon receptor (AhR). Concentrations of anti-inflammatory metabolites, ILA and IAA, were significantly lower in plasma samples of MS patients experiencing acute clinical relapse compared to patients in remission (as shown in Fig. 4b). Additional evidence for the immunomodulatory role of gut microbiota comes from the significant correlation between CSF isobutyrate levels and acute clinical relapse (as illustrated in Fig. 4d). Isobutyrate, a microbial degradation product of valine, is the only short-chain fatty acid known to antagonize β-naphtoflavone-induced AhR activation.^70^ Increased plasma levels of formate in MS patients suffering from acute relapse (as shown in Fig. 4d) may be another indicator of dysbiosis.^71^ Formate dehydrogenase is essential for microbial metabolism in both aerobic and anaerobic environments, maintaining cellular redox balance and energy metabolism by oxidizing formate to carbon dioxide while transferring electrons to an electron acceptor.^72^ In conclusion, shifts in gut microbiota composition may contribute to acute clinical relapse in MS patients through decreased biosynthesis of agonistic and increased production of AhR antagonists.

Unlike during a clinical relapse phase, patients with radiographic disease activity (Gd+) showed higher proportions of IgD^+^IgM^+^CXCR4^+^CD62L^+^ naïve B cells (B7) in the CSF. This population expressed high levels of the trafficking molecule CD62L (L-selectin) and the chemokine receptor CXCR4, known for mediating leukocyte trafficking to sites of inflammation.^73,74^ We also detected a significant increase in plasma and CSF levels of NfL in Gd+ pwMS, particularly those experiencing acute relapse. Notably, no significant changes in NfL expression levels (both in plasma and CSF) were observed between untreated MS and non-MS patients, reinforcing the idea that NfL is not a diagnostic marker for MS but is a broader indicator of ongoing neuroinflammation and axonal damage, as previously reported.^75^ Interestingly, we found a positive correlation between increased levels of CSF NfL and the proportion of CXCR3^+^Tbet^+^CD11c^+^CD38^lo^CD62L^-^ (B6 and B9, see Fig. 1i and 3) atypical memory B cells and CXCR3^+^Tbet^lo^CD11c^-^CD38^+^CD62L^+^ (B10) activated B cells in the CSF of MS patients. These findings suggest that B cell subsets with migratory capacity may be involve in axonal damage. Previous studies have demonstrated that CD11c^+^ B cells can exhibit high potential for both pro-inflammatory and regulatory cytokine production.^76^ The role of these cells in MS is becoming increasingly recognized, as their capacity for antigen presentation and plasma cell differentiation may contribute to disease progression. However, in vivo functional characterization of CSF B cells in human patients remains technically challenging.

Overall, our findings provide key insights into the distinct roles of various B cell subsets in the CSF during clinical and radiographic disease activity in MS. These roles are potentially associated with or consequential to specific molecular factors in the CSF, including metabolites and soluble proteins. These data may have significant implications for diagnosing atypical MS cases and developing targeted therapies that focus on specific B cell populations or molecular factors regulating B cell survival and differentiation. Given the multifactorial nature of MS, characterized by dysregulation of soluble markers, metabolic pathways and cellular processes, the analysis of high-dimensional or multi-omics data and marker clusters (rather than focusing of one single candidate) may provide valuable tools for diagnosis and prognosis. This approach could be particularly useful in differentiating between acute relapse and symptomatic worsening due to other known factors, such as the Uhthoff phenomenon, especially in case without evidence of new T2-weighted lesions on MRI. The absence of new T2-weighted lesions during relapses raises questions about alternative primary mechanistic drivers of disease activity, potentially indicating a more progressive form with reduced inflammation. This observation suggests potential variations in MS disease processes that warrant further investigation. Future studies are necessary to elucidate the clinical utility of these findings in diagnosis, treatment monitoring, and therapy development. Such research may lead to the identification of assessable biomarkers that could be valuable in challenging clinical scenarios.

## Online content

Any methods, additional references, Nature Portfolio reporting summaries, source data, extended data, supplementary information, acknowledgements, peer review information; details of author contributions and competing interests; and statements of data and code availability are available at https://doi.org/10.1038/s41586-024-07516-8.

## Data Availability

Source data are provided with this paper as Source Data file.

## ACKNOWLEDGEMENTS

We would also like to acknowledge the assistance of the BIH Cytometry Core Facility (BIH at Charité – Universitätsmedizin Berlin, Germany). This study was in part funded by F. Hoffmann-La Roche Ltd as part of Integrative Neuroscience Collaborations Network (INTONATE, MOGAD-Precision). M. W. received the PhD scholarship from the Chinese Scholarship Council (CSC). C.B. was funded by the Deutsche Forschungsgemeinschaft (DFG, the German Research Foundation – Project-ID 259373024 – CRC/TRR 167 (B05) and the BMBF (German Ministry of Education and Research), Foerderkennzeichen 01EJ2202A (TAhRget consortium).

## AUTHOR CONTRIBUTIONS STATEMENT

C.B., C.O., K.R. and F.P. conceived and designed the project. C.B., C.F.Z., D.K., G.G., M.W., and A.D. designed and performed mass cytometry analysis. C.O., M.B., F.R.B., C.F., and H.P. recruited MS and non-MS patients and provided the patients’ meta and clinical data. Q.C. and J.L. normalized and analyzed MRI data. B.U. and M.P. performed data and statistical analysis of UK Biobank samples. M.T., C.S., P.J.O. and W.G. performed metabolomics including lipidomics analysis. Ch.B., J.P. and P.S. were responsible for clinical data management and biobanking. C.B., C.F.Z., G.J., C.R., H.R., S.S., R.P., P.J.O., W.G. and K.R. performed data and statistical analysis as well as interpretation of the results. C.B., C.F.Z., C.O., M.W., P.J.O., W.G., J.L., F.P. and K.R. wrote the manuscript.

## ONLINE METHODS

### Ethics statement

The study was approved by the ethics committee of the Charité–Universitätsmedizin Berlin (Ethikkommission der Charité–Universitätsmedizin Berlin; registration number EA1/386/20) in accordance with the Declaration of Helsinki of 1964 and its later amendments. All study participants provided informed consent before any study-related procedures were undertaken and did not receive a compensation.

For the UKB Olink dataset, UKB is a large-scale, population-based cohort with deep genetic and phenotypic data with the full cohort consisting of approximately 500,000 participants. The participants were recruited across centres in United Kingdom and were aged 40 to 69 years at the time of recruitment. Detailed information is available at https://biobank.ndph.ox.ac.uk/showcase/. Ethics approval for the UKB study was obtained from the North West Centre for Research Ethics Committee as a Research tissue biobank (REC reference 11/NW/0382) and all participants provided informed consent. We used the subset of individuals from UKB where both genotype and proteomic measurements were available after excluding ancestry outliers or samples which have failed genomic or proteomics QC (n=43,240). This research has been conducted using the UK Biobank Resource under Application Number 44448.

### Patients’ characteristics

Between 17 December 2020 and 1^st^ June 2023 66 patients with MS, (52 with relapsing-remitting MS (RRMS), 8 with primary progressive MS (PPMS), 3 with secondary progressive MS (SPMS) and 3 with clinically isolated syndrome (CIS)) were included in this prospective observational single-center study at the clinic and outpatient clinic for neurology, Charité-Universitätsmedizin Berlin. 55 patients have newly been diagnosed with MS, respectively CIS and were therapy naïve, whereas 11 patients had confirmed diagnosis for years and/or provided various previous immunotherapy or CIS not suspected for MS. These were excluded from further analysis.

MS patients were further categorized according to treatment status, untreated MS patients (n=55) included those patients who had never received disease modifying therapies (DMT) and did not obtain steroids the day of or at least one day before lumbar puncture. Among the untreated MS patients, 22 suffered from clinical relapse (acute) and 24 patients showed stable course of the disease (stable). Steroid administration at least one day before lumbar puncture occurred in 9 patients with relapse (Acute + Cortisone). Disease duration was defined as interval from onset of symptoms to the timepoint of lumbar puncture. The severity of MS was assessed by a neurological examination performed by an experienced neurologist and evaluated by the Expanded disability status scale (EDSS). We further assessed radiographic disease activity in 25 of the untreated MS patients by systematic assessment of magnetic resonance imaging (MRI) and further differentiated the patients with and without contrast enhancing lesions (Gd^-^ and Gd^+^) respectively. Secondly within each group (Gd^-^ and Gd^+^) patients with acute and stable courses of the disease were assessed separately.

128 patients, 59 women and 69 men, served as controls (non-MS). Diagnoses comprised long-COVID (n = 24), Alzheimer’s disease (n = 46), vascular dementia (n= 2), frontotemporal dementia (n = 2), subjective cognitive impairment (n = 11), mild cognitive impairment (n = 9), epilepsy (n = 3), CBD and Parkinson’s disease (n = 2), neurosyphilis (n = 2), autoimmune encephalitis/-depression (n = 3), cerebellar ataxia (n = 1), controls (n =5), depression (n = 2), leukoencephalopathy (n = 8), MOGAD (n= 2), myelitis (n=2), CIDP after neuroborreliosis (n = 1), somatoform disorder (n = 1), optic neuritis (n = 1), and suspected inflammatory CNS disease (n=1).

Detailed analyses of clinical, demographic and CSF finding are provided in Fig. 1a and Supplementary Tables 1 and 2.

## CyTOF analysis

### Sample processing for CyTOF-based profiling

It is important to note that only CSF samples with erythrocytes cell counts ≤ 500 µl were raised for analysis. Blood and CSF samples were kept on ice (4 °C) before and during the sample preparation, which was performed within 30 min after CSF punction or blood drawing.

Whole blood (EDTA) and CSF cells were fixed in SmartTube Proteomic Stabilizer as described in the user manual and stored at -80°C until CyTOF analysis^77–79^.

### Intracellular barcoding for mass cytometry

After fixation with proteomic stabilizer, whole blood samples were thawed in Thaw/Lyse buffer and subsequently stained with premade combinations of six different palladium isotopes: ^102^Pd, ^104^Pd, ^105^Pd, ^106^Pd, ^108^Pd and ^110^Pd (Cell-ID 20-plex Pd Barcoding Kit, StandardBio). This multiplexing kit applies a 6-choose-3 barcoding scheme that results in 20 different combinations of three Pd isotopes. After 30 min staining (at room temperature), individual samples were washed twice with cell staining buffer (0.5% bovine serum albumin in PBS, containing 2 mM EDTA). All samples were pooled together, washed, and further stained with antibodies.

### Antibodies

Anti-human antibodies (**Supplementary Table 3** for Panel A & **Supplementary Table 4** for Panel B) were purchased either pre-conjugated to metal isotopes (StandardBio) or from commercial suppliers in purified form and conjugated in house using the MaxPar X8 kit (StandardBio) according to the manufacturer’s protocol.

### Surface and intracellular staining

After cell barcoding, washing, and pelleting, the combined samples were re-suspended in 90 μl of antibody cocktail against surface markers and incubated for 30 min at 4 °C. Then, cells were washed twice with cell staining buffer and incubated overnight in 2% methanol-free formaldehyde solution (FA). For intracellular staining, the stained (non-stimulated) cells were subsequently washed once with staining buffer and once with permeabilization buffer (eBioscience). The samples were then stained with 100 μl of the antibody cocktails against intracellular molecules (**Supplementary Tables 3** and **4**) in permeabilization buffer for 30 min at room temperature. Cells were subsequently washed twice with staining buffer, then re-suspended in 1 ml iridium intercalator solution (Fluidigm), and incubated for 30 min at room temperature. Next, the samples were washed twice with cell staining buffer. Cells were pelleted and kept at 4 °C until CyTOF measurement.

### Mass cytometry data processing and analysis

As described previously^77^, nucleated single intact cells were manually gated according to the signals of DNA intercalators ^191^Ir/^193^Ir and event length. For de-barcoding, Boolean gating was used to deconvolute individual samples according to the barcode combination. All de-barcoded samples were then exported as individual FCS files for further analysis. Each FCS file was cleaned and compensated for signal spillover using R package *CATALYST*^80^, transformed with arcsinh transformation (scale factor 5) and batch correction was implemented with a quantile normalization method to minimize batch effects^81^ prior to data analysis. Before performing clustering analyses, CSF cells were pre-gated as follows: from Panel A, CD19^+^ B cells were isolated, and from Panel B, CD19^-^CD3^+^ T cells, along with CD3^-^ myeloid, NK, and B cells (MNB), were extracted (see Supplementary Fig. 1a). For whole blood samples, the gating strategy was as follows: from Panel A, CD3^-^CD19^+^ B cells were selected, and from Panel B, cPARP^-^CD66b^+^ granulocytes, cPARP^-^CD3^+^ T cells, and cPARP^-^CD3^-^CD66b^-^CD14^-/+^ MNK cells were pre-gated using FlowJo. (Supplementary Fig.2). For further clustering analysis we used previously described scripts and workflows^82^. Only samples with >10 cells were considered for the downstream data analysis in the case of CSF samples. For unsupervised cell population identification, we performed cell clustering with the *FlowSOM* and *ConsensusClusterPlus* algorithms using selected markers in each panel (Supplementary Table 5 and 6).

From an initial clustering of CSF cells using Panel A we selected 5 clusters that exhibited higher expression of CD19 and CD20 and subsequently performed sub-clustering with 18 meta- clusters based on the expression of 36 markers including CD19, CD20, CD49d, HLADR, CD38, CCR4, CD24, IgM, CD1c, CXCR4, CXCR3, CD62L, IgA, CD27, CD138, IgGK, CHI3L1, CD11b, IgD, CD28, IgGL, CD28, CD25, Ki67, Tbet, PD1, CTLA4, CD45RO, TNF, KLRG1, CRTH2, CD34, CD3, CD8, TCRgd and CD123. Clustering of CSF cells using Panel B, for the MNB cells we identified 18 meta-clusters based on the expression of 36 markers including CD47, HLADR, CD56, CD66b, CD69, CD95, CD11c, CD14, CD16, CD68, P2Y12, CD33, CD206, CD226, cPARP, CCR7, CD19, CCR4, CD4, CD3, CD141, OPN, CD127, TIGIT, CTLA4, IL6, ICOS, LAG3, TNF, CRTH2, MIP1beta, CXCR4, IL1beta, CD56, CD161 and CD8. As for the T cells we identified 18 meta-clusters based on the expression of, CD3, CD4, CD8, CD47, CRTH2, CTLA4, HLADR, ICOS, TIGIT, CCR4, CD161, CD66b, CD16, CD14, OPN, CD69, CD68, CD11c, CD33, cPARP, CD226 and CCR7.

In Panel A for whole blood samples, the B cells were clustered based on the expression of 36 markers including CD20, C19, HLADR, IgM, CD1c,CD38, CXCR3, CCR4, CD49d, IgA, IgGK, IgGL, IgD, CD138, Tbet, CD28, CD27, CD24, CD25, Ki67, CHI3L1, KLRG1, CD11c, CD11b, CXCR4, CD62L, PD1, CTLA4, CHI3L1, CD45RO, TCRgd, TNF, CD3, CD123, CRTH2 and CD8 and 18-meta clusters were identified.

In panel B for whole blood samples, the T cells compartment was clustered based on the expression of 13 markers, including CD3, CD4, CD127, CCR7, HLADR, CD226, CCR4, CTLA4, ICOS, CD69, LAG3, CD8, CRTH2 and TNF. Eighteen meta clusters were identified within T cell, as well as for myeloid and NK cell compartment and granulocytes compartment. For myeloid and NK cells we used 35 markers, including CD11c, CD68, HLADR, CCR7, CCR4, CD127, CD141, CD33, CD95, CD14, CD64, CD206, CD226, OPN, CD69, CD16, IL6, CD161, TIGIT and IL1beta. For the granulocytes compartment we identified 16 meta-clusters based on the expression of 13 markers including CD95, CD68, CD66b, CD16, CD64, CD14, HLADR, CCR4, CD33, CXCR4, CD161, IL1beta and CD69.

The number of metaclusters used for further analysis was identified based on the delta area plots (which asses the “natural” number of clusters that best fits the complexity of the data) together with visual inspection on the phenotypic heatmap with the aim to select a cluster number with consistent phenotypes that would also allow to explore small populations. For dimensionality-reduction visualization we generated UMAP representations using all markers as input and down-sampled to a maximum of 1000 cells per sample. Differences of cluster abundances between groups were analyzed using linear mixed models (limma package) including age as covariate. The Benjamini-Hochberg FDR correction was applied to account for multiple testing, with statistical significance defined at a 5% FDR cutoff.

### NULISA protein quantification

Protein concentrations from CSF samples (MS = 44, non-MS = 62) were measured with the Alamara NULISAseq™ 120-plx CNS disease panel, which mainly targets neurodegenerative disease-related protein markers and immune response-related cytokines/chemokines, and the NULISAseq™ 250-plex Inflammation Panel, mainly targeting immune response-related cytokines/chemokines. The Alamar NULISA immunoassays utilizes a pair of antibodies for each target, one capture antibody conjugated with a partially double-stranded DNA containing a poly-A tail and a target-specific molecular identifier and a second detection antibody conjugated with another partially double-stranded DNA containing a biotin group and a matching target-specific barcode. The barcode sequence is then quantified using next generation sequencing.

Protein concentrations are normalized with internal and inter-plate controls and log2 transformed, these values are then defined as NULISA Protein Quantification (NPQ) units, which were used for subsequent analysis. We excluded one datapoint that did not pass the internal quality control. Additionally, four samples showed values below the limit of detection (LOD); however, as advised by the company, these samples were included in the analysis, as they did not significantly impact the results. Differentially expressed proteins (DEPs) between groups were identified using linear mixed models with the limma R package (version 3.58.1) adjusted for age. Benjamini-Hochberg false discovery rate (FDR) correction was applied to account for multiple testing, and statistical significance was defined using a 5% FDR cutoff. We then used the differentially expressed proteins (DEPs) to perform protein-protein interaction (PPI) network analysis on the STRING database (https://cn.string-db.org/) via the stringApp 2.0 in Cytoscape (version 3.10.01), with a confidence score of 0,4 and no additional interactors.

### Olink proteomics

Proteomic profiling was performed on CSF (MS = 57, non-MS = 19) samples using the Olink Explore 3072 platform, capturing 3,072 unique proteins targeted by 3,072 assays, which included the following panels: Cardiometabolic, Cardiometabolic II, Inflammation, Inflammation II, Neurology, Neurology II, Oncology and Oncology II each comprised of 384 unique assays. Olink technology employs proximity extension assays that utilize pairs of antibodies conjugated to complimentary oligonucleotides. Upon binding to their respective target proteins, hybridization between probes enables amplification, followed by relative quantification through next generation sequencing.

Normalized protein expression (NPX) values are generated by normalizing to the extension control, applying a log2 transformation, and further normalizing to the plate controls. NPX units were used for all downstream analysis. All datapoints that did not meet internal quality control standards were excluded. Subsequent analysis was performed using the OlinkAnalyze package for R (version 4.0.1). DEPs between groups were identified using linear mixed effects model with age as a covariate. The Benjamini-Hochberg FDR correction was applied to account for multiple testing, with statistical significance defined at a 5% FDR cutoff. The DEPs were further analyzed for PPI network using the STRING database (https://cn.string-db.org/) via the stringApp 2.0 in Cytoscape (version 3.10.01), with a confidence score of 0,4 and no additional interactors.

The UKB proteomic measurements were conducted for plasma samples of ∼54,000 participants using the antibody-based Olink technology, Explore 3072 platform. Further details about antibody-based proteomic measurements and QC have been described elsewhere, including the exclusion of samples due to poor quality and selective measurements with assay warnings (Gadd et al. 2024). After initial quality control checks (e.g., principal component analysis), we did further filter to individuals with at least 50% valid protein measurements. Notably, UKB provided proteomic data with values flagged as assay warnings by Olink as ‘NA’, which meant that each protein measurement had differing numbers of valid values and we decided not to impute values for the purpose of this study. We included a total of 2919 unique protein targets in our analysis. To evaluate the association between plasma protein levels and multiple sclerosis, we fitted linear regression models for each protein assay with the formula: Protein level (NPX) ∼ disease_type + age + sex + bmi + age^2^ + age:sex + age^2^:sex + technical_covariates (The technical covariates include fasting time, time and month of blood sampling, sample age). Bonferroni correction was used to account for multiple testing. We compared the Olink measurements of UK Biobank participants with prevalent multiple sclerosis (n=369) to those with other neurodegenerative disorders (n=182, including Parkinson’s disease, Alzheimer’s disease and anterior horn cell disease), other autoimmune disorders (n=922, including systemic lupus erythematosus, ankylosing spondylitis, coeliac disease, Crohn’s disease, ulcerative colitis, rheumatoid arthritis, and psoriasis), and controls (n=49214). We excluded participants with incident diagnoses of any of the listed conditions.

## NMR spectroscopy

For metabolomic and lipidomic analysis, NMR measurements of 286.72 mL of unfiltered EDTA-plasma specimens were mixed with 286.72 μl of Bruker plasma IVDr NMR buffer (Bruker BioSpin GmbH, Rheinstetten). To this end a SamplePro (Bruker) sample preparation robot was employed. NMR experiments were carried out on a 600 MHz Bruker Avance III (Bruker BioSpin GmbH) employing a triple-resonance (1H, 13C 15N, 2H lock) cryogenic probe equipped with z-gradients and an automatic cooled sample changer. For each sample a 1D 1H NOESY, a 2D 1H JRES, a 1D 1H CPMG, and a 1D 1H diffusion experiment was acquired at 310 K as specified in the Bruker IVDr methods. This experimental data was transferred to the Bruker analysis server, where absolute concentrations of 41 metabolites and 134 lipid parameters were automatically determined.

For NMR measurements of CSF samples 300.00 mL of CSF were mixed with 300 mL NMR buffer (Bruker BioSpin GmbH, Rheinstetten). As an internal reference standard that is not prone to protein binding 20 mL of 240.00 mmol/L formic acid (FA) were added. Employing the 600 MHz NMR spectrometer described above, for each sample a 1D Carr-Purcell-Meiboom-Gill (CPMG) experiment was performed at 298 K. Raw NMR data were semiautomatically processed employing TopSpin 4.0.7 (Bruker BioSpin GmbH). Spectra were quantified with Chenomx 8.6 (Chenomx Inc.,Edmonton, AB, Canada) resulting in the quantification of a total of 25 compounds in each spectrum.

Tables of resulting metabolite concentrations and lipid parameters from plasma and metabolite concentrations from CSF were imported into the statistical-analysis software R version 4.2.3 (Development Core Team, R. R: A Language and Environment for Statistical Computing. (4.2.3). 2023. Vienna, Austria, R Foundation for Statistical Computing). For the computation of ANOVAs the R limma package was used (Smyth2004).

## MRI

### Image acquisition

All participants underwent contrast-enhanced MRI of the brain using clinical 1.5T- (17 patients) or 3.0T- (14 patients) MRI scanners. The MRI protocol for each patient varies from each other, especially the specific parameters, including T1-weighted imaging (T1WI), T2-weighted imaging (T2WI) and fluid attenuated inversion recovery (FLAIR) and contrast-enhanced MRI. A gadolinium-based contrast agent was administered according to the patient’s total body weight.

### Imaging process and analysis

Upon brain extraction, each sequence was co-registered with T1WI (ANTs, version 2.4.3).^80^ Subsequently, all co-registered images were transformed to atlas space (T1-ICBM152 2009c).^81^ Segmentation of lateral ventricles (LV) from the MNI-ICBM152 CerebrA atlas,^82^ were transformed from atlas to subject space, thereby generating a binary LV mask for each subject. LV segmentation was visually assessed and corrected where necessary. White matter lesion (WML) and contrast-enhancing lesions (CEL) were manually segmented on co-registered T2WI and Gd-T1WI images using ITK-SNAP^83^ by two experienced MRI technicians (more than 10 years of experience) and then revised by two neuroradiologists (Q.C. and J.L., 6 and 7 years of experience) in consensus. The total volume and number of lesions were calculated using fslmaths and cluster from FSL (version 6.0). The normalized LV volume was calculated as ratio of LV volume and total intracranial volume (TIV).

### Multi-omics data integration

To identify multi-omic signatures we used the Data Integration Analysis for Biomarkers Discovery using Latent cOmponents (DIABLO), a multi-block sparse partial least squares discriminant analysis (sPLS-DA) method from the mixOmics package. DIABLO uses supervised discriminant analysis while maximizing the correlated information between different datasets. The design matrix was set with a weight of 0.1 between datasets and 1 between each dataset and the outcome variable, to prioritize discrimination. We manually selected between 5 and 10 variables from each dataset to reduce computational time, note that we only selected the number of variables while the model selected the individual variables. For visualization, we plotted the individuals in the two-dimensional space defined by the weighted average from the first two principal components.

### Statistical analysis

Unless otherwise stated, statistical analysis was performed using GraphPad Prism (version 8.0.2). Continuous variables were expressed as mean and standard deviation with or without range. Mann-Whitney U-tests were used to analyze differences in detected number of cells, cluster frequencies and differences in metabolite concentrations. Correlations between CSF immune cell frequencies and Olink or Nulisa DEPs were calculated with Spearman correlation at a confidence level of 0.95, for plotting the circlize R package was used and for visualization, only links above a p-value threshold of <0.05 and a correlation cut off of ≤-0.4 or ≥0.4 were selected. Significances were presented as *p<0.05; **p<0.01 and ***p<0.001.

## Data availability

Source data are provided with this paper as Source Data file.

## Code availability

Codes used for CyTOF data analysis in this study are previously published by Crowell H et al. 2022 and available on https://github.com [https://github.com/HelenaLC/CATALYST]. Code used for analysis of Olink data has been previously published and is available on https://github.com/Olink-Proteomics/OlinkRPackage.

