## Supplementary Figures for "Comprehensive cerebrospinal fluid analysis indicates key roles for B cells in multiple sclerosis"

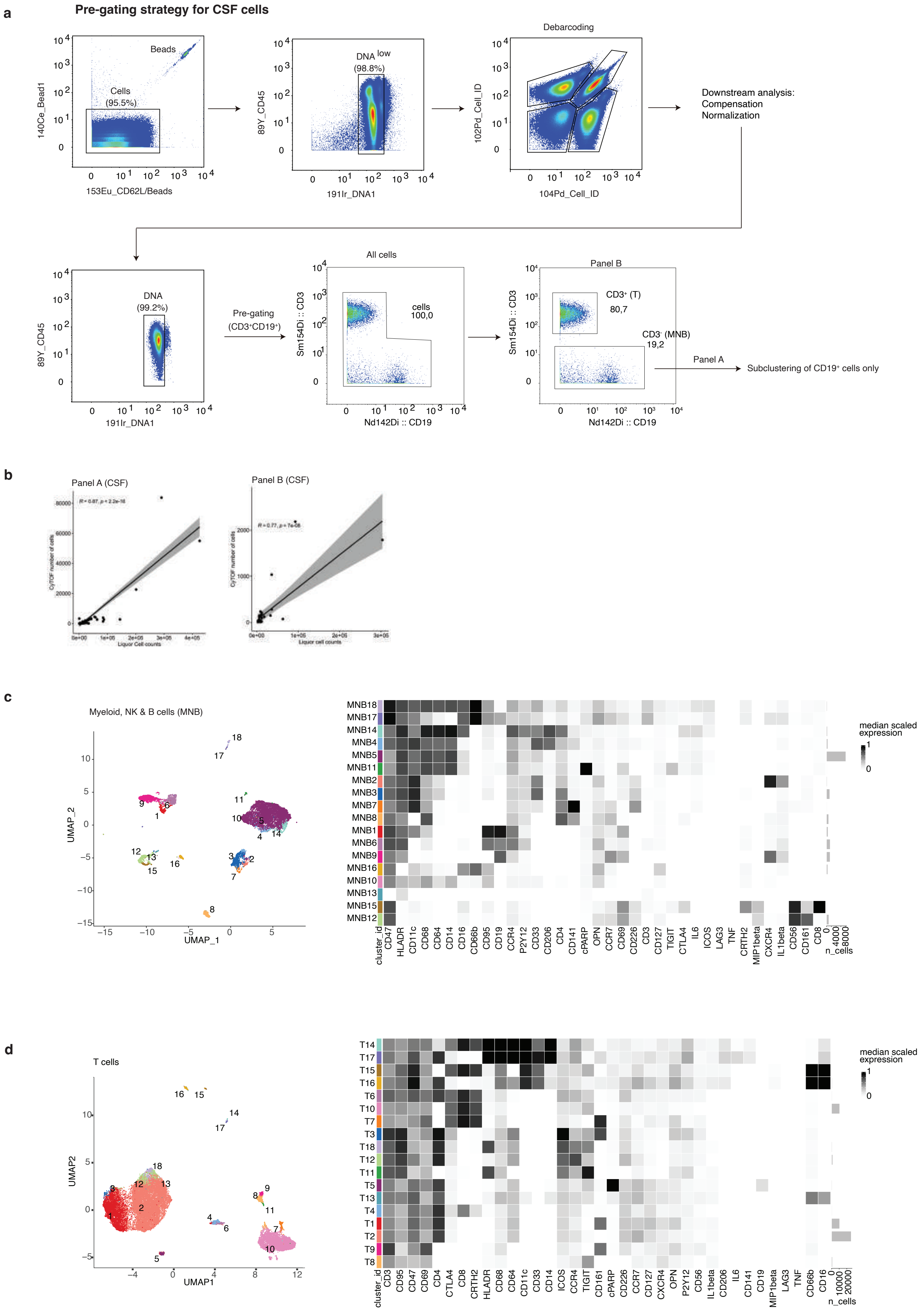


Extended Data Figure 1

**Extended Data Figure 1. Pre-gating strategy for CSF cells. a,** Gating strategy showing removal of cell debris, doublets and beads, followed by debarcoding. After this step, files are transferred to R for compensation of signal spillover and signal normalization between batches. All files are then imported into FlowJo for pre-gating of $CD3^+$ (T cells) or $CD3^-$ (myeloid, NK and B cells, referred to as, MNB) for Panel B or for subclustering of B cells from the $CD3^-$ population in the case of Panel A. **b,** Spearman's correlation between routine cell counts and the number of cells detected with CyTOF, regression line with 95% confidence interval. **c, d,** Unsupervised clustering analysis of MNB and T cells, respectively. UMAP plots are shown in the left panel, where each dot represents one cell, and phenotypic heatmaps displaying median-scaled marker expression are shown in the right panel.

**Pre-gating strategy for whole blood cells**

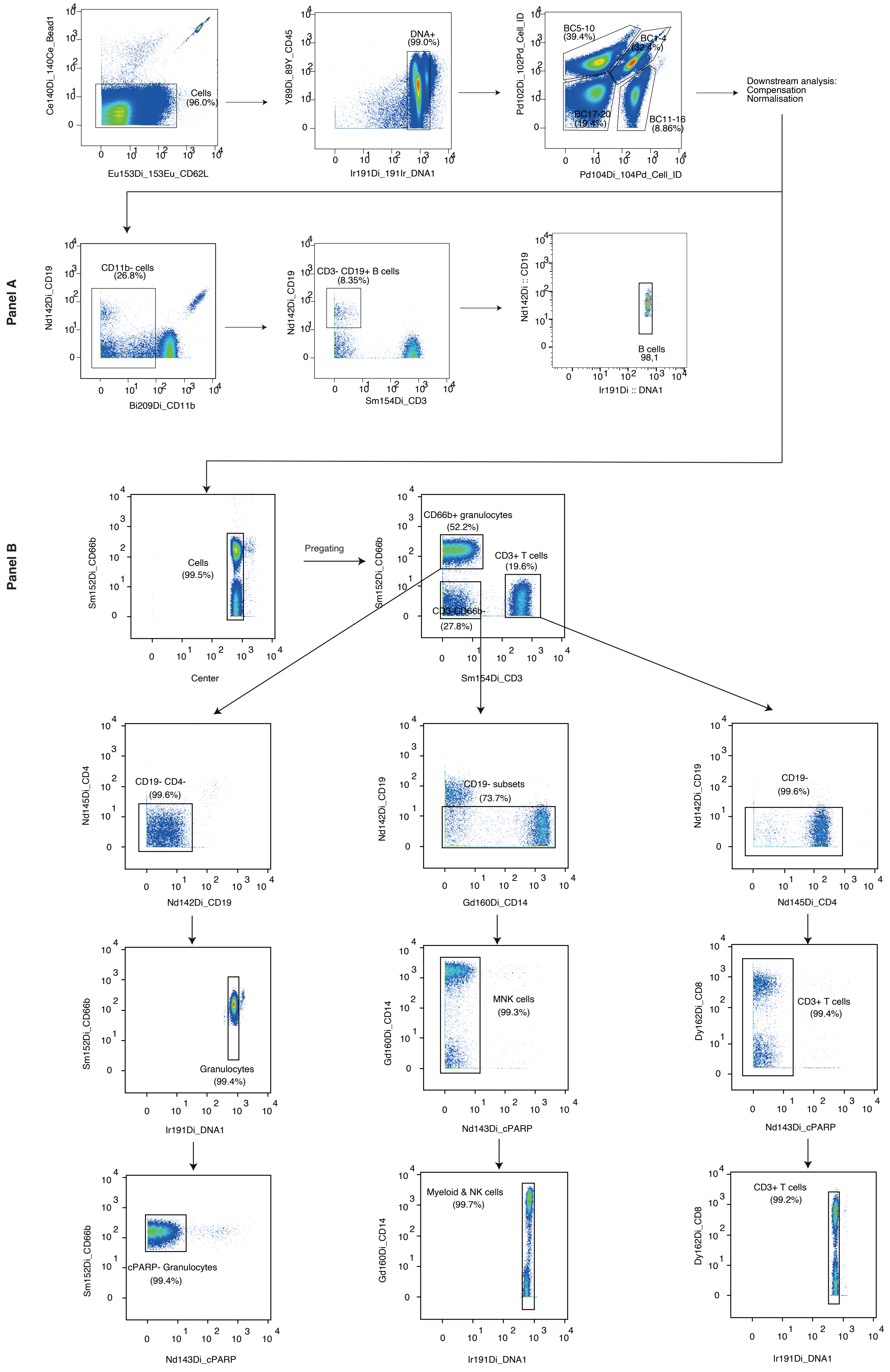


Extended Data Figure 2

**Extended Data Figure 2. Pre-gating strategy for WB cells. a,** Gating strategy showing removal of cell debris, doublets and beads, followed by debarcoding, after which files are transferred to R for compensation of signal spillover and normalization of signal intensity between batches. Gating strategies for specific selection of B cells from Panel A and granulocytes, MNK cells and T cells from Panel B are depicted in density plots.

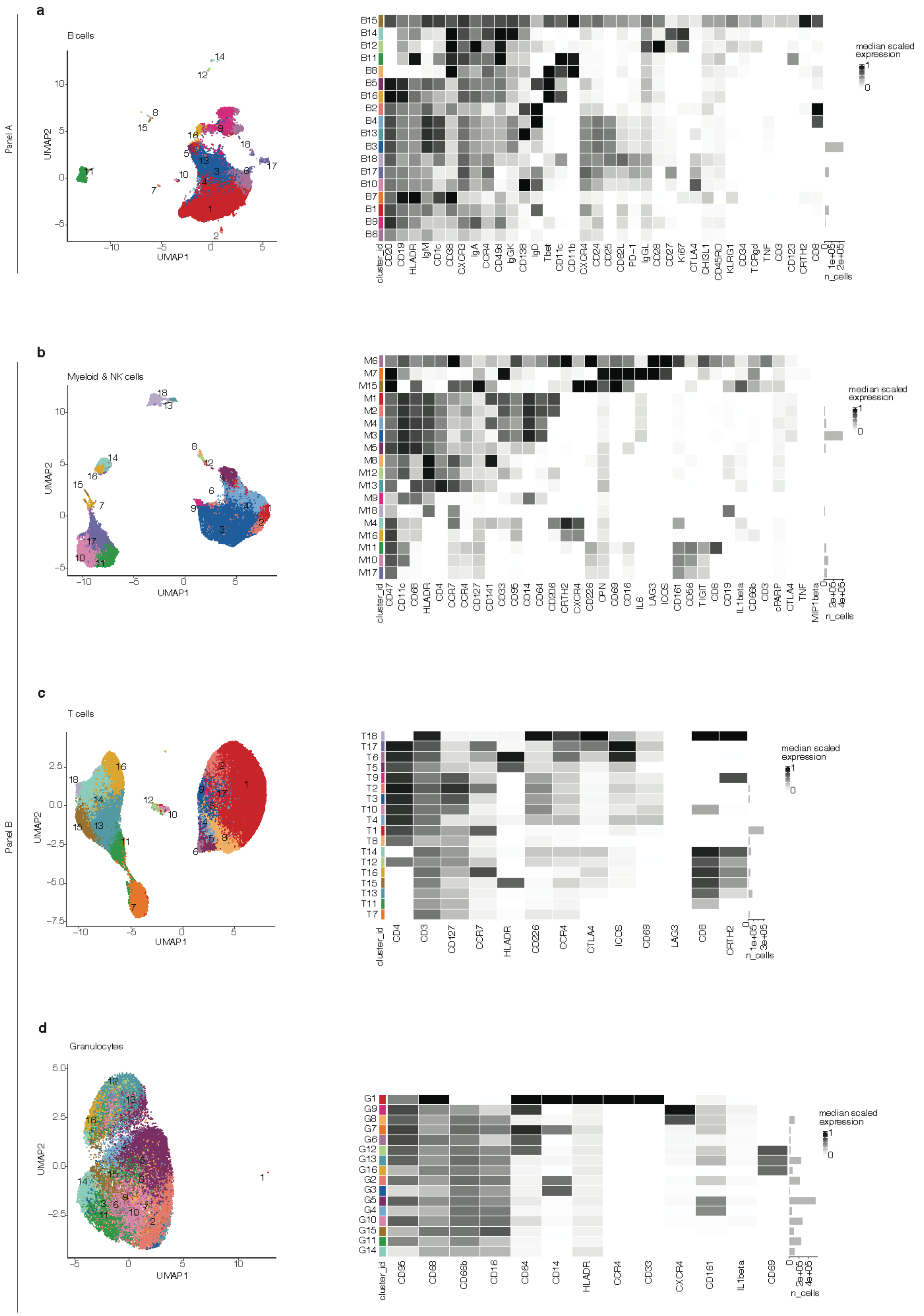


Extended Data Figure 3

**Extended Data Figure 3. Unsupervised clustering of WB populations.** Unsupervised clustering was performed to detect subpopulations of B cells from Panel A (**a**). From Panel B, myeloid and NK cells (**b**), T cells (**c**) and granulocytes (**d**). UMAP plots colored by cluster ID are shown on the left, with corresponding phenotypic heatmaps of median scaled expression presented on the right.

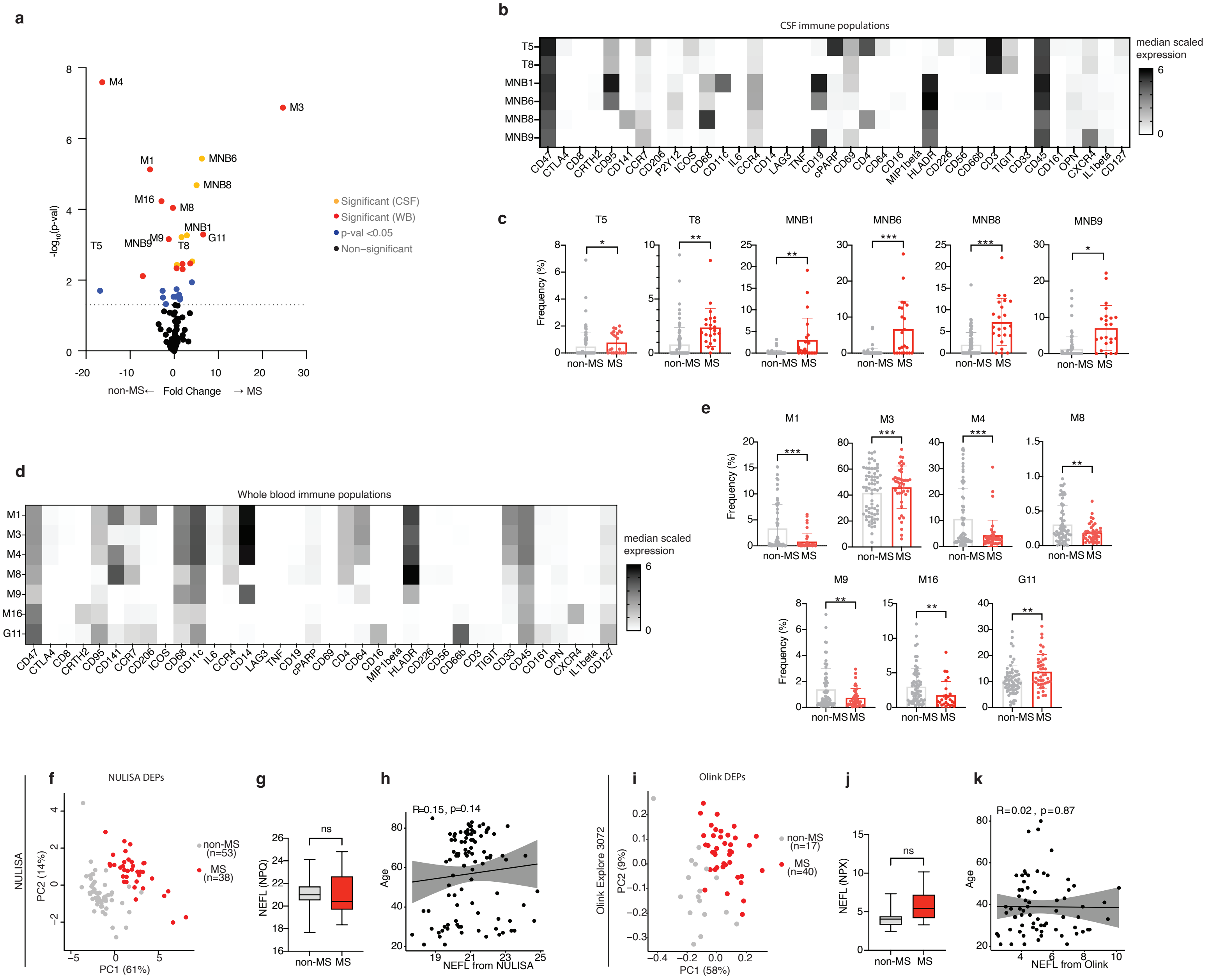

Extended Data Figure 4

**Extended Data Figure 4. Changes in WB and CSF populations between MS and non-MS patients. a,** Volcano plot displaying differences in frequency of WB and CSF immune clusters measured with CyTOF between non-MS (n=60) and untreated MS (n=23) patients, with exclusion of B cells from the CSF. Analysis was conducted using linear mixed effects model adjusted for age. Clusters with statistically significant difference between groups after correction for multiple testing (Benjamini & Hochberg) are colored in red (WB) or yellow (CSF); populations with *p*-value <0.05 colored in blue, others are in black. Each dot represents one cluster and horizontal dashed line represents a *p*-value threshold of 0.05. **b**,**d**, Phenotypic heatmaps of CSF (**b**) and WB (**d**) immune populations that are differentially abundant between groups. **c**,**e**, Bar plots showing detailed frequency distribution of CSF (**c**) and WB (**e**) immune clusters differentially abundant between groups. Data are shown as mean ± SD. **f,i,** PCA plot of the NULISA and Olink DEPs, each dot represents one patient. **g,j,** Boxplot showing NfL levels in the CSF of non-MS and MS patients measured with Olink Explore and NULISA platforms respectively. **h**,**k**, Scatter plots showing correlation between NfL levels as measured with Olink (**h**) or NULISA (**k**) platforms and age of the participants, assessed with Spearman's correlation coefficient, regression line with 95% confidence interval. . *$p < 0.05$, **$p < 0.01$, ***$p < 0.001$.

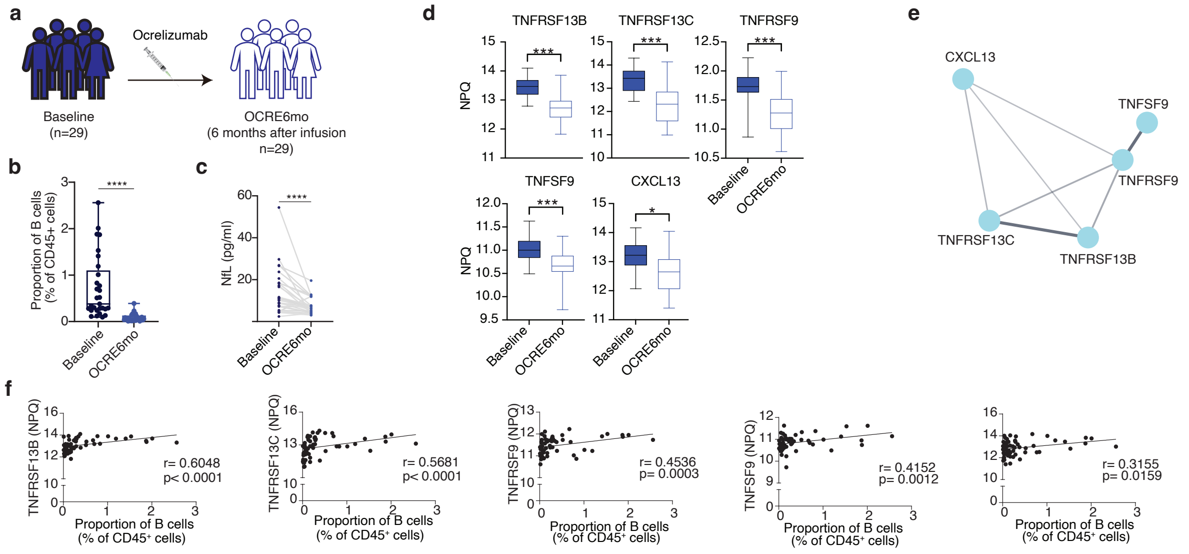


Extended Data Figure 5

**Extended Data Figure 5. Molecular changes in plasma following ocrelizumab treatment in MS patients. a,** Schematic depiction of ocrelizumab (OCRE) treated cohort. **b,** Proportion of whole blood B cells between baseline and 6 months after ocrelizumab treatment (OCRE6mo). Data are shown as mean ± SD. **c,** NfL levels in the plasma of MS patients at baseline and 6 months after ocrelizumab treatment. **d,** NPQ expression levels for the DEPs in plasma between baseline and after 6 months of ocrelizumab treatment measured with NULISA. Data are shown as mean ± SD. **k**, Protein interaction network of the DEPs, obtained using the STRING databased and visualized with Cytoscape. Only proteins with direct interactions are shown. **f,** Spearman's correlations between proportion of B cells in WB and plasma DEPs measured with NULISA. **p* < 0.05, ***p* < 0.01, ****p* < 0.001.

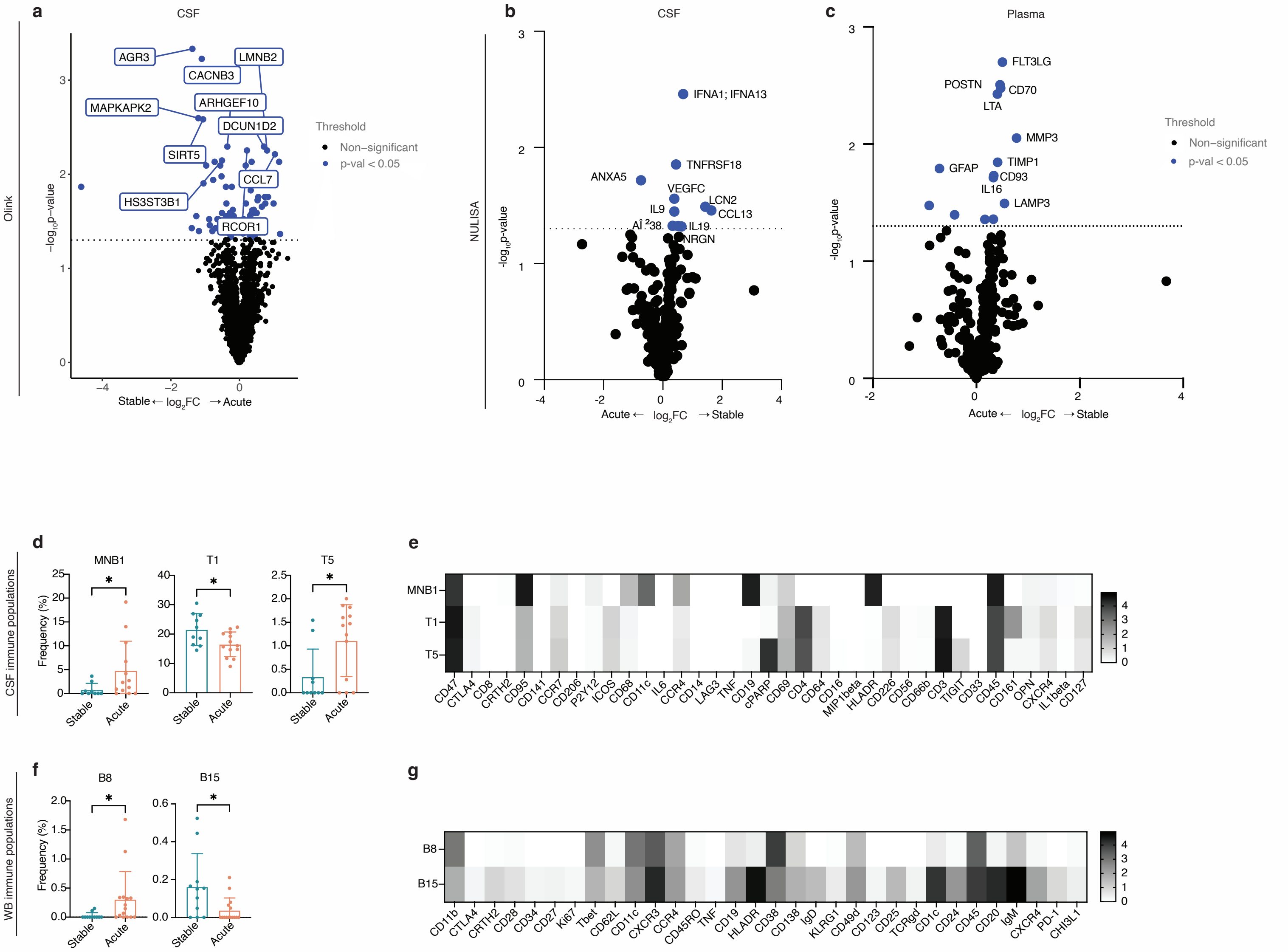


Extended Data Figure 6

**Extended Data Figure 6. Changes in cellular and molecular markers between untreated MS patients with stable and acute clinical disease course. a, b, c,** Volcano plots displaying differences in protein concentrations of markers analyzed with NULISA (stable, n= 17; acute, n=19) from CSF and plasma and Olink (stable, n=21; acute, n=18) from CSF samples. Analysis was conducted using linear mixed effects model adjusted for age. Markers with statistical significance difference between groups after correction for multiple testing (Benjamini & Hochberg) are colored in red; markers with *p*-value <0.05 are colored in blue, others are in black. Each dot represents one protein and horizontal dashed line represents a *p*-value threshold of 0.05. **d,f** Barplots of differentially abundant CSF (stable, n=10; acute, n=13) (**d**) and WB (stable, n=11; acute, n=13) (**f**) immune populations between untreated stable and acute MS patients and corresponding phenotypic heatmaps displaying median-scaled marker expression. Statistical analysis in **d** and **f** using two-tailed Mann–Whitney U-test. Data are shown as mean ± SD. (**e, g**). $^*p < 0.05$, $^{**}p < 0.01$, $^{***}p < 0.001$.

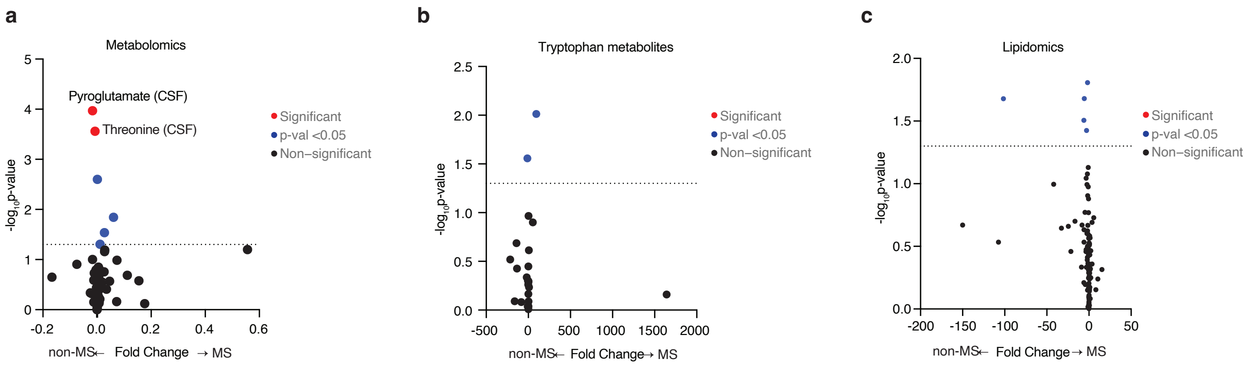


Extended Data Figure 7

**Extended Data Figure 7. a, b, c,** Volcano plots showing differences in metabolomics (**m**), tryptophan metabolites (**n**) and lipidomics (**o**) between non-MS and MS patients. Analysis was conducted using linear mixed effects model adjusted for age. Statistical significance difference between groups after correction for multiple testing (Benjamini & Hochberg) are colored in red; values with *p*-value <0.05 colored in blue, others are in black. Each dot represents one metabolite or lipid species, and horizontal dashed line represents a *p*-value threshold of 0.05. Metabolomics (non-MS, n=91; MS, n=36), tryptophan metabolites (non-MS, n=85; MS, n=30), lipidomics (non-MS, n=83; MS, n=34).

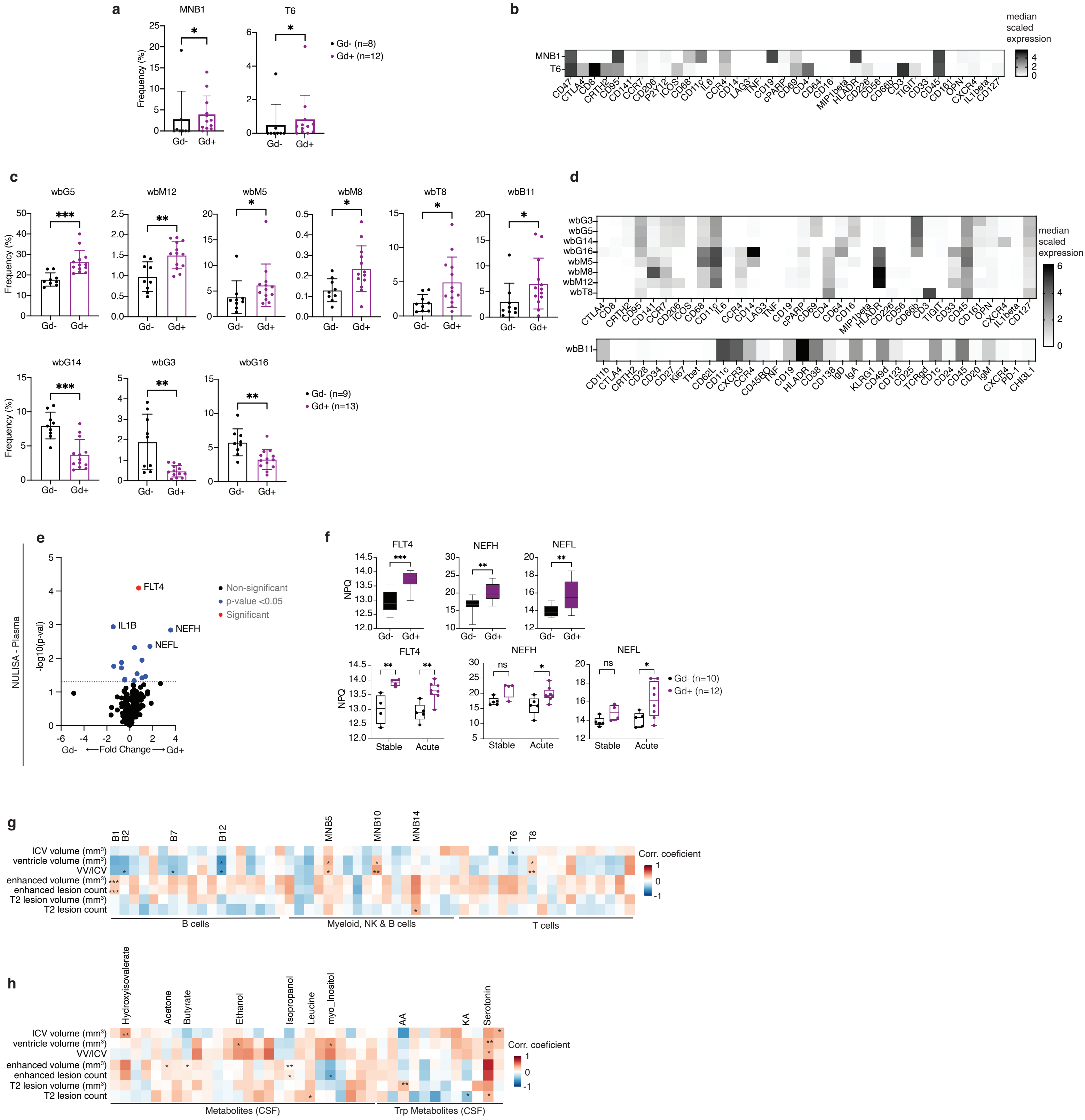


Extedend Data Figure 8

**Extended Data Figure 8. Changes in cellular and molecular markers between untreated MS patients with and without Gd-enhanced lesions. a,c** Barplots of differentially abundant CSF (**a**) and WB (**c**) immune populations between Gd- and Gd+ untreated MS patients and corresponding phenotypic heatmaps of median-scaled expression (**b** and **d**). **e,** Volcano plots displaying differences in protein concentrations of markers analyzed with NULISA platforms from plasma samples (Gd-, n=10; Gd+, n=12). Analysis was conducted using linear mixed effects model adjusted for age. Markers with *p*-value <0.05 are colored in blue, others are in black. Each dot represents one protein, and horizontal dashed line represents a *p*-value threshold of 0.05. **d,** Box plots of significantly different proteins between Gd- and Gd+ untreated MS patients, further subdivided into stable and acute patients. **e,** Correlations between MRI variables and CSF immune populations. Data are shown as mean ± SD **f,** Correlations between MRI variables and metabolomics. Calculated with Spearman's correlation. $*p < 0.05$, $**p < 0.01$, $***p < 0.001$.
